# Longitudinal Associations Between White Matter Microstructure and Psychiatric Symptoms in Adolescence

**DOI:** 10.1101/2022.08.27.22279298

**Authors:** Lorenza Dall’Aglio, Bing Xu, Henning Tiemeier, Ryan L. Muetzel

## Abstract

**Objective:** Associations between psychiatric problems and white matter (WM) microstructure have been reported in childhood and adolescence. Yet, a deeper understanding of this relation has been hampered by a dearth of well-powered longitudinal studies and a lack of explicit examination of the bidirectional associations between brain and behavior. We investigated the temporal directionality of WM microstructure and psychiatric symptom associations from late childhood to early adolescence.

**Methods:** In this observational study, we leveraged the world’s largest single- and multi-site cohorts of neurodevelopment: the Generation R and Adolescent Brain Cognitive Development Studies (total n scans = 11,400). We assessed psychiatric symptoms with the Child Behavioral Checklist as broad-band Internalizing and Externalizing scales, and as syndrome scales (e.g., Anxious/Depressed). We quantified WM with Diffusion Tensor Imaging, globally and at a tract level. We used cross-lagged panel models to test bidirectional associations of global and specific measures of psychopathology and WM microstructure, meta-analyzed results across cohorts, and used linear mixed-effects models for validation.

**Results:** We did not identify any robust longitudinal associations of global WM microstructure with internalizing or externalizing problems across cohorts (confirmatory analyses). We observed similar findings for longitudinal associations between tract-based microstructure with internalizing and externalizing symptoms, and for global WM microstructure with specific syndromes (exploratory analyses).

**Conclusion:** Uni- or bi-directionality of longitudinal associations between WM and psychiatric symptoms was not robustly identified. We propose several explanations for these findings, including interindividual differences, the use of longitudinal approaches, and smaller effects than expected.

## INTRODUCTION

Neural correlates of psychiatric problems have been widely studied, including internalizing and externalizing symptoms in childhood and adolescence^1–4^. Internalizing problems indicate a child’s internal distress (e.g., depression) and generally increase during adolescence^5^. Externalizing problems refer to overt behaviors creating conflict with others (e.g., aggression) and are prominent in childhood but tend to stabilize or decrease over time. Both internalizing and externalizing problems are important predictors of adverse outcomes and psychopathology in adulthood^6,7^. Identifying their neurobiological substrates is, therefore, of great importance, but it has been challenging, with findings being inconsistent across studies.

Associations have been reported between internalizing and externalizing symptoms with measures of white matter (WM) microstructure, such as fractional anisotropy (FA) and mean diffusivity (MD). FA and MD respectively indicate the directional preference and magnitude of water diffusion in the brain^8^, with FA increasing and MD decreasing in typically developing adolescents^9^. It has been suggested, with inconsistencies^10^, that children with higher psychiatric symptoms present delayed neurodevelopmental trajectories^11–14^. This might imply that normative changes in behavioral and WM microstructural development have not yet occurred or are less prominent in children at a given age. Cross-sectional studies have generally shown that higher psychiatric symptoms are associated with lower FA and higher MD^15–17^. However, our understanding of the longitudinal relation between neuro- and behavioral development is limited by a dearth of well-powered longitudinal studies with repeated measurements^18^ and by the overlooking of how brain and behavior relate to each other over time.

Dissecting these temporal relations is paramount for understanding the role of WM microstructure in the development and continuation of psychiatric problems. Typically, it is assumed that neurobiology determines psychiatric problems^19^. While mental illness undoubtedly has a neurobiological substrate, it has been suggested that the opposite temporal relation may also occur. Psychiatric symptoms can shape brain development^20^. In fact, there are several biological and social elements of psychiatric problems, such as anxiety, which are related to neurodevelopment, including heightened physiological arousal, altered sleep patterns, and fewer social interactions^21,22^. The possibility of bidirectional relations between brain connectivity and psychological functioning is termed *probabilistic epigenesis* and is a key assumption of the interactive specialization framework^23^. Evidence for probabilistic epigenesis in child psychiatry would hold major implications by highlighting the importance of timely interventions before potential deviations in neurodevelopment may occur. Yet, the temporal dynamics of neurobiological and psychiatric associations remain largely overlooked.

A few studies with repeated measurements have emerged on the topic: some highlighted that brain structural anatomy and connectivity precede changes in psychiatric symptoms, and others found the opposite temporal relation. For instance, Lin et al.^24^, in a clinical sample of children to young adults (*N* = 56), showed that frontoparietal structural connectivity predicted changes in autism severity, but not the other way around. Albaugh et al.^25^ highlighted that brain structure during adolescence determined changes in hyperactivity and inattention symptoms in young adulthood (*N* = 976) – the opposite temporal relation was not examined. In contrast, Muetzel et al.^26^, in a younger sub-cohort of one of the present population-based studies (Generation R (GenR), *N* = 845), found that psychiatric symptoms in early childhood predicted brain structural and microstructural changes in late childhood. Conversely, brain structure was related to psychiatric symptoms cross-sectionally but did not predict symptom changes over time. Moreover, further studies showed that anxiety, depression, and inattention symptoms predicted the rate of cortical and/or microstructural development^11,12^.

While this literature offers some insight into the potential temporal relations between neurobiology and psychiatric problems, the current understanding of the topic is limited in several ways. First, only two studies explicitly tested a bidirectional relation^24,26^. Second, previous research often focused on specific psychiatric problems. However, unlike the broad constructs of internalizing and externalizing symptoms, some psychiatric problems are not as stable through childhood and into adolescence (e.g., anxiety)^5^. Such instability complicates longitudinal studies of brain development, meaning that a global approach to measuring psychiatric symptoms may be preferable. Third, prior literature generally sampled youth with a wide age range. This could dilute effects and reduce the power to detect changes in associations across a specific developmental period. Lastly, the transition from childhood to adolescence, a key developmental window, characterized by marked brain plasticity and the emergence and exacerbation of many psychiatric problems^27,28^, remains largely uncharted in longitudinal studies of psychopathology and neurodevelopment.

Here, we examined the temporal relations of global WM microstructure and internalizing and externalizing problems in the transition from late childhood to early adolescence. We hypothesized that lower global FA and higher global MD in childhood would predict larger increases in internalizing problems and smaller decreases in externalizing problems from childhood to adolescence. We also hypothesized that higher internalizing and externalizing problems in childhood would predict smaller increases in global FA and decreases in global MD from childhood to adolescence. To address our aims, we leveraged the world’s largest multi- and single-site pediatric cohorts of neurodevelopment: the Adolescent Brain Cognitive Development (ABCD) and GenR Study cohorts, for a total of 5,700 participants with repeated MRI and behavioral assessments (n total scans = 11,400). These independent studies have similar measures sampled at overlapping participant ages within a narrow time period. Our hypotheses and analysis plans were preregistered at https://osf.io/pny92.

## METHODS

### Participants

This longitudinal observational study is embedded within two existing pediatric population-based cohorts: the ABCD and GenR Studies. ABCD has been conducted across 21 study sites within the U.S., where pre-adolescents have been sampled and will be followed up until adulthood^29^. GenR has been conducted at one study site in Rotterdam, the Netherlands, and it spans from fetal life until late adolescence^30^. Ethical approval was received from the institutional review boards of the University of California (San Diego) and of each of the 21 data collection sites for ABCD, and the Medical Ethics Committee of Erasmus MC, University Medical Centre (Rotterdam) for GenR. Informed consent or assent has been received from the included participants. More information on the samples is available elsewhere^29,30^.

We used data from the assessments at ages 10 and 12 for ABCD and 10 and 14 years for GenR. These assessment waves were chosen to maximize comparability across cohorts. We included children with data on internalizing and externalizing problems and DTI global measures (FA, MD) at both time points.

For ABCD, children were excluded based on the recommended guidelines from the cohort team (v.4.0, DOI: 10.15154/1523041), and clinically relevant incidental findings (**Supplementary Text**). Post-hoc, children who changed site from baseline to follow-up were excluded due to statistical issues with modeling site (*n* = 53, **Supplementary Text**), and one child was additionally excluded due to invalid data in the DTI measures (i.e., global FA value = 0.98, **Supplementary Text**).

For GenR, children were excluded if they had clinically relevant incidental findings or poor quality images (e.g., due to artifacts). Due to differences in scan sequence, a small proportion of children was also excluded (*n* =89).

For each set of remaining siblings or twins in either cohort, one was randomly included. The final sample constituted 4,605 children from ABCD and 1,095 from GenR, for a total of 5,700 children and 11,400 imaging sessions (**Supplementary Figure 1** for the inclusion flowchart).

### Measures

#### Image Acquisition

Children were scanned using multiple 3T scanners across sites for ABCD (General Electric (GE) 750, Philips, Siemens Prisma) and on a single scanner for GenR (GE 750w) (Casey et al., 2018; Muetzel et al., 2017) (**Supplementary Text** for more details).

#### Image pre-processing

In ABCD, pre-processing was performed by the ABCD Data Analysis, Informatics & Resource Centre and was previously described in the literature^31^ and the ABCD Curated Annual Release notes (v.4.0 DOI: 10.15154/1523041). In GenR, DTI images were pre-processed through the FMRIB Software Library (FSL) (v. 6.0.2)^32^. This included removal of non-brain tissue, eddy-current induced artifact correction^33^, and volume realignment for simple head motion translations and rotations. A weighted least-squares method was used to fit the diffusion tensor at each voxel. Commonly used scalar metrics were then obtained (e.g., FA, MD).

#### WM probabilistic tractography

In ABCD, WM tracts were identified with AtlasTrack, a probabilistic atlas-based approach automatizing tract segmentation^34^. Each MRI image was nonlinearly registered to the atlas with the use of discrete cosine transforms and the obtained diffusion orientations were then compared to the atlas fiber orientations. Voxels that mostly included cerebral spinal fluid or grey matter were excluded. For each tract identified with AtlasTrack, weighted averages for DTI measures were calculated. For bilateral tracts (i.e., having separate left and right hemisphere measures), values were averaged.

In GenR, probabilistic fiber tractography was run on each child’s native space diffusion data by leveraging the FSL plugin “AutoPtx”, which automatically identified connectivity distributions for commonly reported fiber bundles. A library of seed, target, and exclusion masks was leveraged to generate the connectivity distributions by applying a nonlinear transformation from the FMRIB FA map to each subject’s native space. Normalization of connectivity distributions was performed using the number of successful seed-to-target attempts. Where voxels seemed unlikely to be part of the true distribution, they were removed by thresholding^35^. The average values for the FA and MD of each WM tract (per hemisphere) were obtained after weighting the voxels based on the connectivity distributions. Left and right hemisphere values were averaged per tract. Finally, to obtain global connectivity measures, we averaged all FA or MD values across 10 commonly used tracts and weighted this average based on the volume of each tract.

#### Image Quality Control

In ABCD, QC was performed both at the pre-processing and post-processing stages with manual and automated checks and is described elsewhere^31^ (**Supplementary Text**). In GenR, manual and automatic QC was performed for DTI images. For the manual QC, visual inspection was conducted based on the sum-of-squares error of the tensor calculation and the tract reconstructions. For the automated QC, translation and rotation motion parameters for the eddy tool were additionally used to exclude data with excessive motion (**Supplementary Text**).

#### Psychiatric symptoms

Psychiatric symptoms were measured using the Child Behavioral Checklist (CBCL) in both cohorts. The CBCL is a parent-reported inventory to measure child psychopathology dimensionally. It is a reliable and valid instrument, generalizable across nationalities and societies^36,37^. The CBCL/6-18 (school-age version) was administered at both assessment waves in each cohort to the mother or primary caregiver. It consists of 112 problem items, rated on a scale from 0 (not true) to 2 (very true or often true), which can be scored on eight narrow-band syndrome scales (e.g., Anxious/Depressed), and broad-band scales such as Internalizing, Externalizing, and Total problems^36^. To preserve natural variation in the data, raw scores were used for both cohorts. Due to non-normal distributions, variables were square-root transformed.

### Covariates

The following covariates were considered in both cohorts to eliminate their potential confounding influence on the results: child age, sex, ethnicity, perceived pubertal status, highest achieved parental education, study site (for ABCD only), age difference in behavioral and MRI assessment (for GenR only). Assessment information can be found in the **Supplementary Text**.

Of note, the names of all the variables described here are specified in **Supplementary Table 1** for reproducibility purposes.

### Statistical Analyses

All analyses were conducted with the R statistical software (version 4.0.3) and all scripts will be publicly available at https://github.com/LorenzaDA/DTI_psychiatry_youth_ABCD-GenR. Any deviation from our preregistered analyses is specified in **Supplementary Table 2**.

#### Confirmatory analyses

To investigate the bidirectionality of WM microstructure – psychiatric symptoms associations, we ran *cross-lagged panel models* (CLPMs). These permit the examination of the associations between two variables with repeated measurements simultaneously. Several coefficients are estimated: lagged effects, covariances (i.e., cross-sectional associations between brain and psychiatric problems), and autoregressive coefficients (i.e., stability over time). Of these, we were interested in the lagged effects, i.e. *(i)* where internalizing/externalizing problems predict *changes* in WM microstructure (by accounting for baseline levels), and *(ii)* where WM microstructure predicts *changes* in internalizing/externalizing problems.

The CLPMs were run in the R *lavaan* package (version 0.6.8)^38^. Four CLPMs were run for each cohort, based on each combination of internalizing and externalizing problems with global FA or MD (**Supplementary Text**). To account for covariate missingness (see **Table 1** for amounts of missing values), full-information maximum likelihood was used within the *lavaan* package. Correction for multiple testing was performed with the False Discovery Rate (FDR) method (Benjamini–Hochberg procedure), which was applied to all tested associations (total n = eight). The hypothesized models were *(i)* compared to competing models to test for model equivalence, which is good practice in structural equation modeling^39^, *(ii)* adapted based on modification indices where model fit was suboptimal according to prespecified criteria (RMSEA ≤ 0.06; SRMR ≤ 0.09; https://osf.io/pny92; **Supplementary Text**). **Figure 1** displays the final models (fit indices in **Supplementary Table 3**). To maximize power and highlight replicable results, estimates for each lagged path were standardized and subsequently meta-analyzed (fixed-effects, weighted by sample size) using the R package *meta* ^40^. An *a priori* power analysis was conducted (see preregistration at https://osf.io/pny92). Overall, we were well-powered (>80%) to detect effect sizes as large or smaller than the ones identified in prior literature on the topic for most tested associations.

**Table 1.**
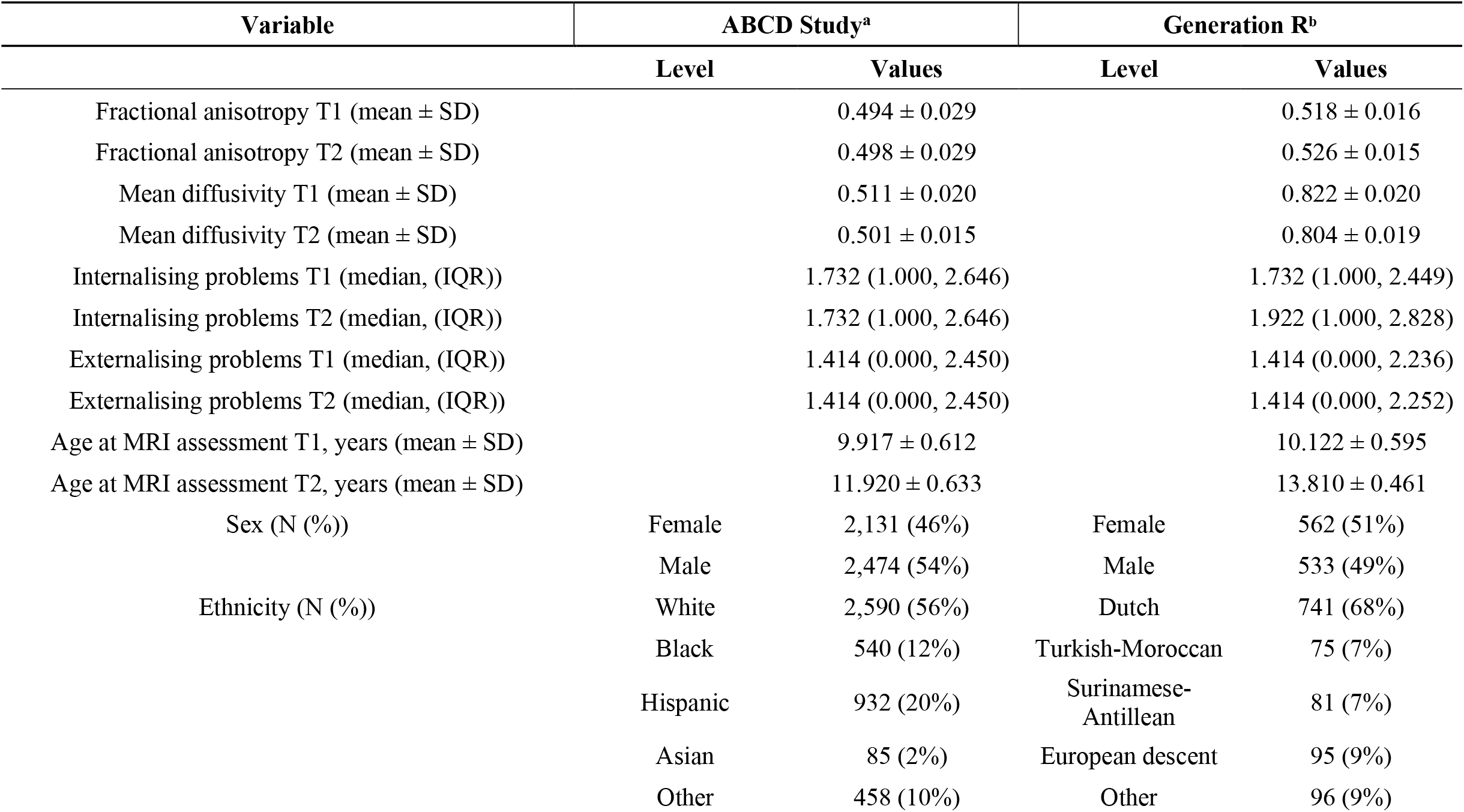
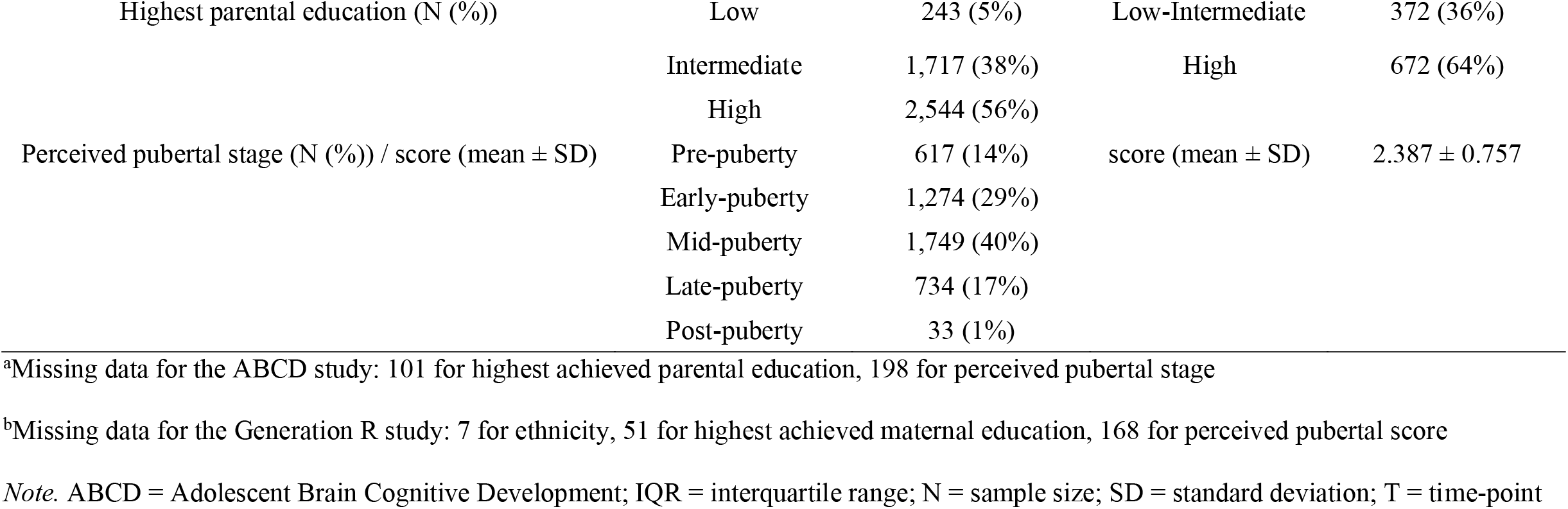
Descriptive statistics of the sample population of the ABCD and Generation R Studies.

| Variable | ABCD Study<sup>a</sup> |  | Generation R<sup>b</sup> |  |
| --- | --- | --- | --- | --- |
|  | Level | Values | Level | Values |
| Fractional anisotropy T1 (mean ± SD) | | 0.494 ± 0.029 | | 0.518 ± 0.016 |
| Fractional anisotropy T2 (mean ± SD) | | 0.498 ± 0.029 | | 0.526 ± 0.015 |
| Mean diffusivity T1 (mean ± SD) | | 0.511 ± 0.020 | | 0.822 ± 0.020 |
| Mean diffusivity T2 (mean ± SD) | | 0.501 ± 0.015 | | 0.804 ± 0.019 |
| Internalising problems T1 (median, (IQR)) |  | 1.732 (1.000, 2.646) |  | 1.732 (1.000, 2.449) |
| Internalising problems T2 (median, (IQR)) |  | 1.732 (1.000, 2.646) |  | 1.922 (1.000, 2.828) |
| Externalising problems T1 (median, (IQR)) |  | 1.414 (0.000, 2.450) |  | 1.414 (0.000, 2.236) |
| Externalising problems T2 (median, (IQR)) |  | 1.414 (0.000, 2.450) |  | 1.414 (0.000, 2.252) |
| Age at MRI assessment T1, years (mean ± SD) | | 9.917 ± 0.612 | | 10.122 ± 0.595 |
| Age at MRI assessment T2, years (mean ± SD) | | 11.920 ± 0.633 | | 13.810 ± 0.461 |
| Sex (N (%)) | Female | 2,131 (46%) | Female | 562 (51%) |
|  | Male | 2,474 (54%) | Male | 533 (49%) |
| Ethnicity (N (%)) | White | 2,590 (56%) | Dutch | 741 (68%) |
|  | Black | 540 (12%) | Turkish-Moroccan | 75 (7%) |
|  | Hispanic | 932 (20%) | Surinamese-Antillean | 81 (7%) |
|  | Asian | 85 (2%) | European descent | 95 (9%) |
|  | Other | 458 (10%) | Other | 96 (9%) |
| Highest parental education (N (%)) | Low | 243 (5%) | Low-Intermediate | 372 (36%) |
|  | Intermediate | 1,717 (38%) | High | 672 (64%) |
|  | High | 2,544 (56%) |  |  |
| Perceived pubertal stage (N (%)) / score (mean ± SD) | Pre-puberty | 617 (14%) | score (mean ± SD) | 2.387 ± 0.757 |
|  | Early-puberty | 1,274 (29%) |  |  |
|  | Mid-puberty | 1,749 (40%) |  |  |
|  | Late-puberty | 734 (17%) |  |  |
|  | Post-puberty | 33 (1%) |  |  |

<sup>a</sup>Missing data for the ABCD study: 101 for highest achieved parental education, 198 for perceived pubertal stage
<sup>b</sup>Missing data for the Generation R study: 7 for ethnicity, 51 for highest achieved maternal education, 168 for perceived pubertal score
*Note.* ABCD = Adolescent Brain Cognitive Development; IQR = interquartile range; N = sample size; SD = standard deviation; T = time-point

**Figure 1.**
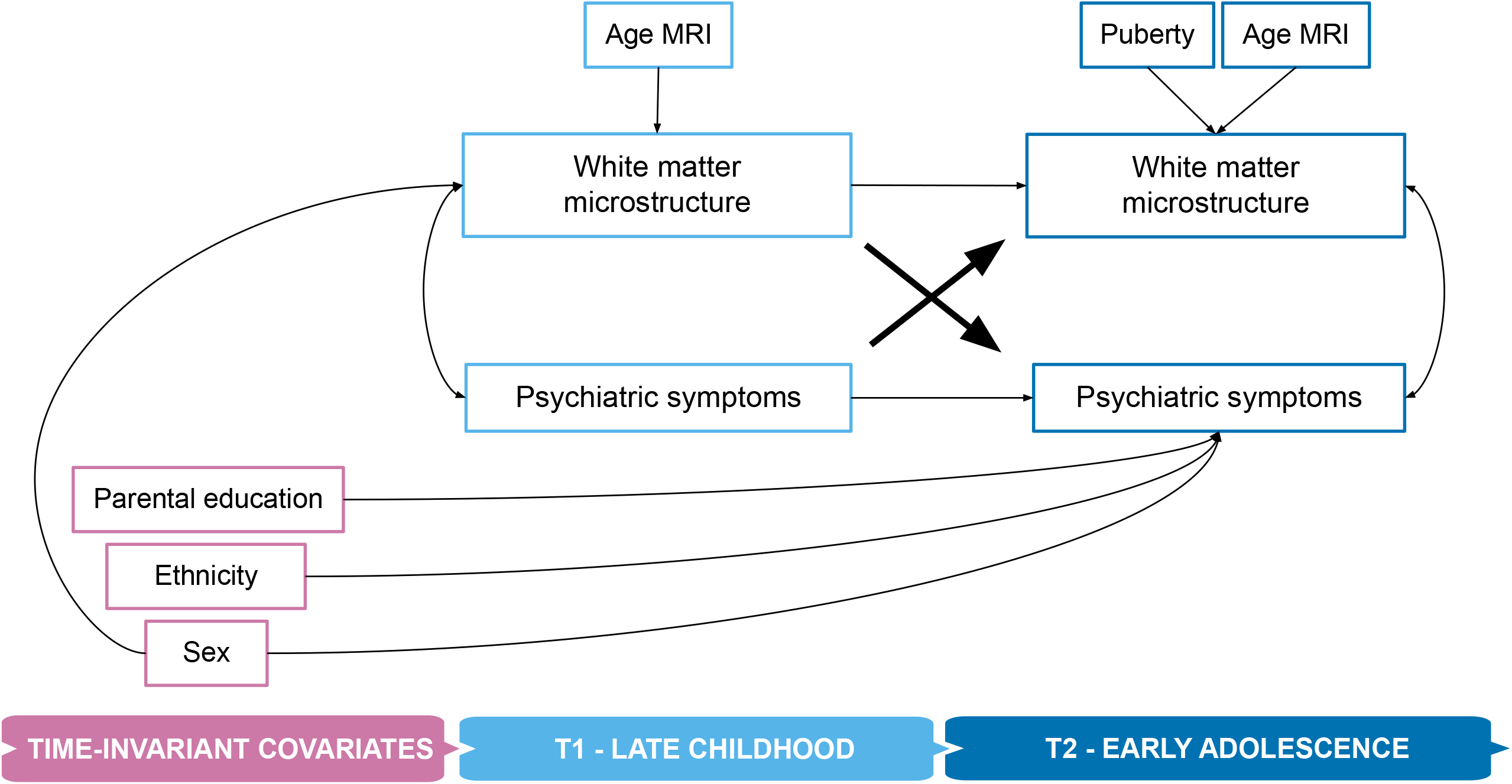
The cross-lagged panel model *Note*. MRI = magnetic resonance imaging; T = time-point The effect of site on white matter microstructure was additionally included for the ABCD Study due to its multi-site structure. The effects of the difference in age at MRI and psychiatric assessment on white matter microstructure were also included for the Generation R Study, as it assessed children’s white matter connectivity and behavioral problems at different child ages. Paths of interest (lagged paths) are bolded and of larger size.

#### Sensitivity Analyses

We conducted sensitivity analyses to evaluate the robustness of the results. Firstly, we examined relations between WM and internalizing/externalizing symptoms using *linear mixed-effects* models (for each lagged path of interest), run with the *lme4* package^41(p4)^ (**Supplementary Text**). Linear mixed-effects models complemented CLPMs by examining intra-individual variability over time (random intercepts).

Secondly, we ran a non-response analysis in GenR, which, being a birth cohort, allows us to quantify potential attrition bias. We compared children invited to the assessments at the age of 10 years (a similar invitation used for prior assessment rounds) with those who participated in both the behavioral and MRI data collections at age 10 and 14 years, based on covariates (sex, puberty, ethnicity, parental education). We used independent samples t-tests and chi-square tests for continuous and categorical covariates, respectively.

#### Exploratory analyses

Several exploratory analyses were conducted to better understand longitudinal relations between WM connectivity and psychiatric symptoms. First, we explored how *specific psychiatric problems* (i.e., the eight CBCL syndrome scales) longitudinally related to global FA and MD with CLPMs (**Supplementary Text**). The same analyses were performed for *general psychopathology*, as measured by the total problem scale of the CBCL, with global FA and MD, and for 10 WM connectivity tracts with internalizing and externalizing problems (**Supplementary Text**). We meta-analyzed results across cohorts for each analysis and corrected for multiple testing (*p*FDR < 0.05) within each set. Finally, we explored whether our main results differed by sex in each cohort, in line with guidelines to bridge the gender data gap (e.g., from agencies like NIH, ERC) (**Supplementary Text**).

## RESULTS

### Sample characteristics

We included 4,605 individuals from ABCD (46% females) and 1,095 from GenR (51% females) (**Table 1**). Children were, on average, 10 years old at the first assessment (T1) for both cohorts, and 12 and 14 years old at the second assessment (T2) for ABCD and GenR, respectively (**Figure 2A&D, Table 1**) (rangeABCD : 9-14 years; rangeGenR : 9-16 years). Children generally showed increases in FA and internalizing problems (GenR only), decreases in MD, and stable levels of externalizing problems over time (**Figure 2B&E; Table 1**). Relations between psychiatric symptoms and WM microstructure are depicted in **Figure 2C&F** for both cohorts (bivariate Pearson correlations).

**Figure 2.**
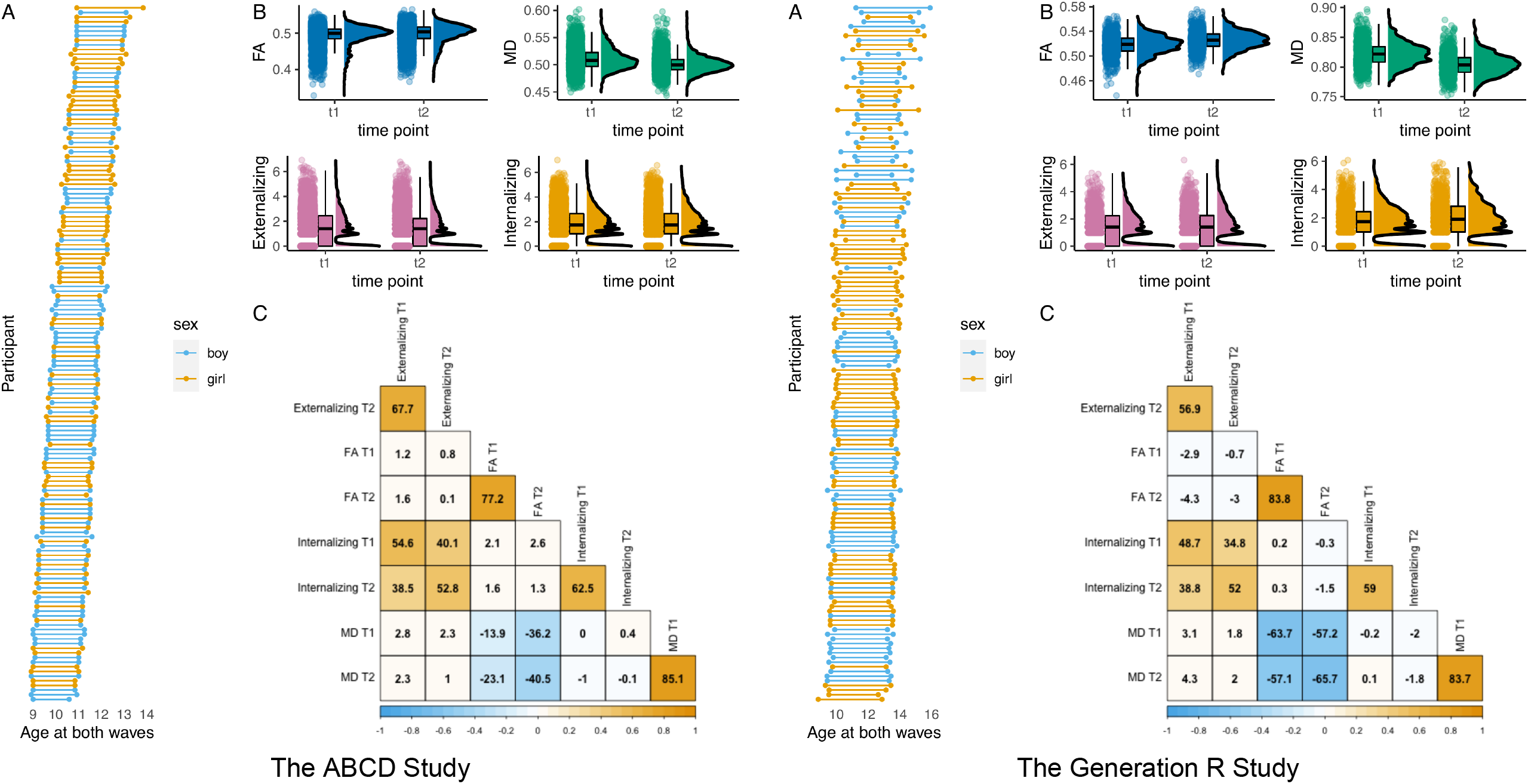
Sample and variable characteristics for the ABCD (left) and Generation R (right) Studies *Note*. On the left, sample characteristics for the ABCD Study are shown. On the right, sample characteristics for the Generation R Study are displayed. **A&D**. Representative subset of participants visualizing age at assessment for time-point 1 and 2, grouped by sex. **B&E**. Variable distribution for FA, MD, externalizing, and internalizing problems across time-point 1 and 2. **C&F**. Correlation plot between FA, MD, externalizing, and internalizing problems over time (Correlation coefficients are scaled by 100 for visualisation purposes). FA = fractional anisotropy; MD = mean diffusivity.

### Confirmatory and sensitivity analyses

We focused on the longitudinal relations between psychiatric problems and global WM microstructure, represented by the lagged paths of the cross-lagged panel models (**Figure 1**, bolded arrows). None of the paths were statistically significant in the meta-analysis or either cohort separately (**Table 2**). The meta-analysis showed modest standardized absolute coefficients for the lagged paths, with a median of 0.009, ranging from 0.000 to 0.013. Standard errors were relatively large (median: 0.010; range: 0.006 – 0.010).

**Table 2.** Confirmatory analyses results from the cross-lagged panel models for the meta-analysis and each study.

|  | Meta-analysis CLPM |  |  | ABCD CLPM |  |  | GenR CLPM |  |  |
| --- | --- | --- | --- | --- | --- | --- | --- | --- | --- |
| Model | Estimate | SE | *p*-value | Estimate | SE | *p*-value | Estimate | SE | *p*-value |
| *Internalizing problems* |  |  |  |  |  |  |  |  |  |
| FA<sub>T1</sub> → Internalizing<sub>T2</sub> | 0.009 | 0.010 | 0.392 | 0.010 | 0.011 | 0.388 | 0.004 | 0.025 | 0.862 |
| MD<sub>T1</sub> → Internalizing<sub>T2</sub> | 0.013 | 0.010 | 0.210 | 0.014 | 0.011 | 0.203 | 0.006 | 0.025 | 0.817 |
| Internalizing<sub>T1</sub> → FA<sub>T2</sub> | -0.005 | 0.006 | 0.372 | -0.006 | 0.006 | 0.319 | 0.002 | 0.017 | 0.888 |
| Internalizing<sub>T1</sub> → MD<sub>T2</sub> | -0.000 | 0.007 | 0.970 | -0.001 | 0.008 | 0.866 | 0.004 | 0.016 | 0.797 |
| *Externalizing problems* |  |  |  |  |  |  |  |  |  |
| FA<sub>T1</sub> → Externalizing<sub>T2</sub> | 0.006 | 0.010 | 0.534 | 0.006 | 0.011 | 0.560 | 0.005 | 0.025 | 0.825 |
| MD<sub>T1</sub> → Externalizing<sub>T2</sub> | 0.005 | 0.010 | 0.621 | 0.006 | 0.011 | 0.590 | 0.000 | 0.025 | 0.998 |
| Externalizing<sub>T1</sub> → FA<sub>T2</sub> | -0.006 | 0.006 | 0.325 | -0.004 | 0.006 | 0.495 | -0.017 | 0.017 | 0.309 |
| Externalizing<sub>T1</sub> → MD<sub>T2</sub> | -0.003 | 0.007 | 0.645 | -0.009 | 0.008 | 0.253 | 0.020 | 0.016 | 0.202 |

*Note.* ABCD = The Adolescent Brain Cognitive Development study; CLPM = cross-lagged panel model; FA = fractional anisotropy; GenR =
The Generation R study; MD = mean diffusivity; SE = standard error; T = time-point

Similar results were found in linear mixed-effects models. A relation between externalizing problems at baseline and changes in MD over time was found in ABCD, but could not be replicated in GenR and did not survive multiple testing correction (**Supplementary Table 4**). The remaining tested models did not show any relation between internalizing/externalizing problems with WM microstructure, in either temporal direction. Further corroborating these results, equivalent model fit was shown with competing models, which had lagged paths between brain and behavior set to 0 (**Supplementary Table 5**). Lastly, a non-response analysis in GenR showed that participants were representative of the full cohort at T1 (**Supplementary Table 6**), suggesting that results were likely robust to attrition.

### Exploratory Analyses

We ran several exploratory analyses to better understand the relation between WM microstructure and psychiatric symptoms in adolescence. First, we tested *specific psychiatric problems* (8 CBCL syndrome scales) with global FA and MD. Results for each cohort and the meta-analysis are shown in **Figures 3-4** (global FA only) and **Supplementary Table 7** (global FA and MD). For brain to behavior, no statistically significant association was observed, nominally or after FDR correction, in the meta-analysis. For behavior to brain, social problems predicted smaller changes in FA over time (meta-analysis estimate = -0.016, SE = 0.006, *p* = 0.005), but did not survive FDR correction (*p*FDR = 0.148). Overall, of the 32 tested lagged paths, none survived multiple testing correction in the meta-analysis. The median absolute effect size was 0.007, with median standard errors of 0.008.

**Figure 3.**
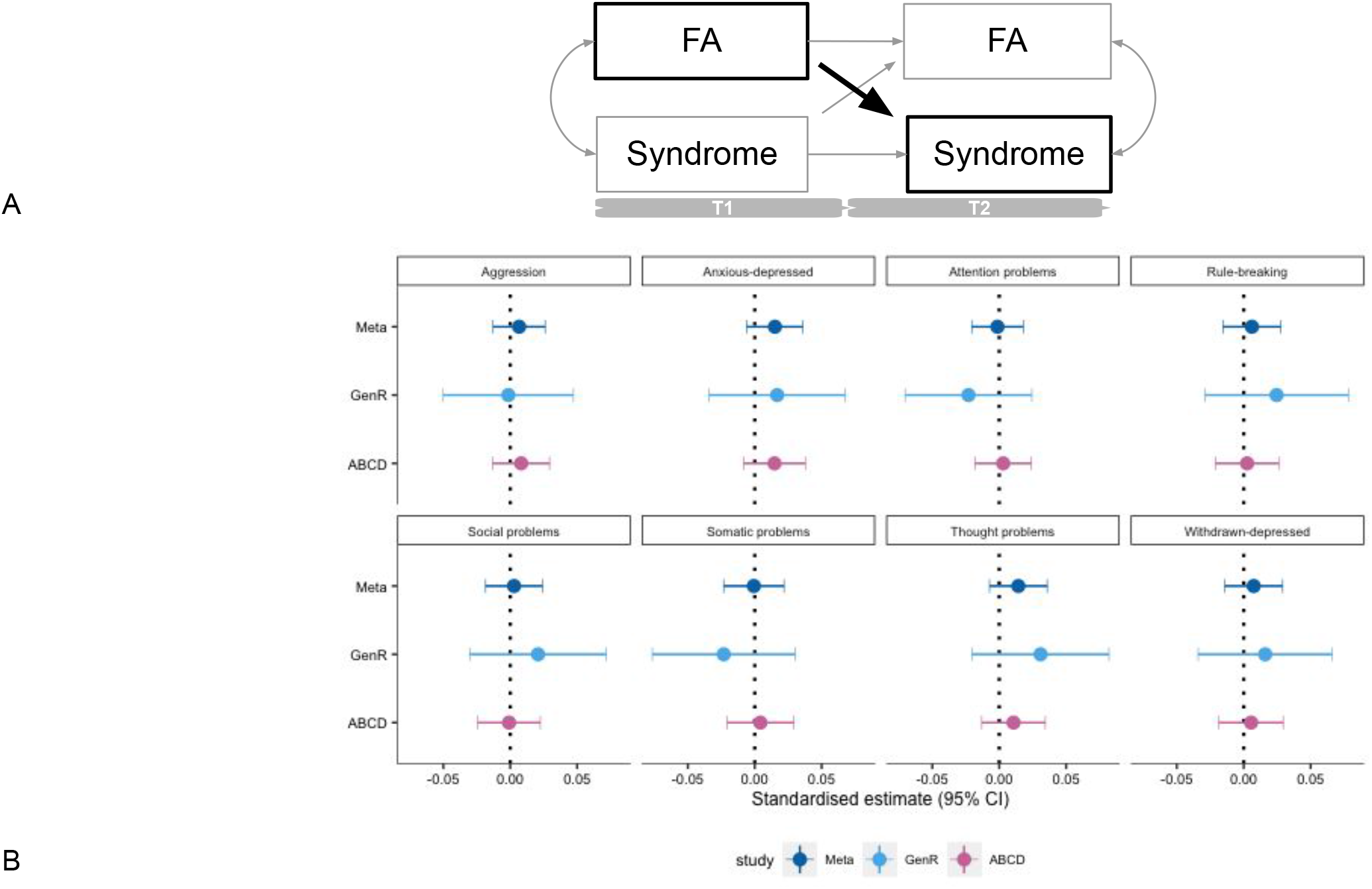
Meta-analysis and single study results for the relation between baseline global FA and changes in psychiatric symptoms (syndrome scales). *Note*. ABCD = The ABCD Study; GenR = The Generation R Study; FA = fractional anisotropy; Meta = meta-analysis; Syndrome = CBCL Syndrome Scale; T = time-point. **A**. Cross-lagged panel model with lagged paths of interest in bold and larger font: global FA at T1 → psychiatric symptoms T2. **B**. Standardized coefficients for the effect of global FA at T1 on changes in the 8 syndromes scales from T1 to T2. Results are grouped by syndrome scale. Coefficients from the meta-analysis and for each cohort are displayed. Positive coefficients indicate higher FA at baseline is related to larger changes in psychiatric symptoms. Negative coefficients point to higher FA at baseline being related to lower changes in psychiatric symptoms.

**Figure 4.**
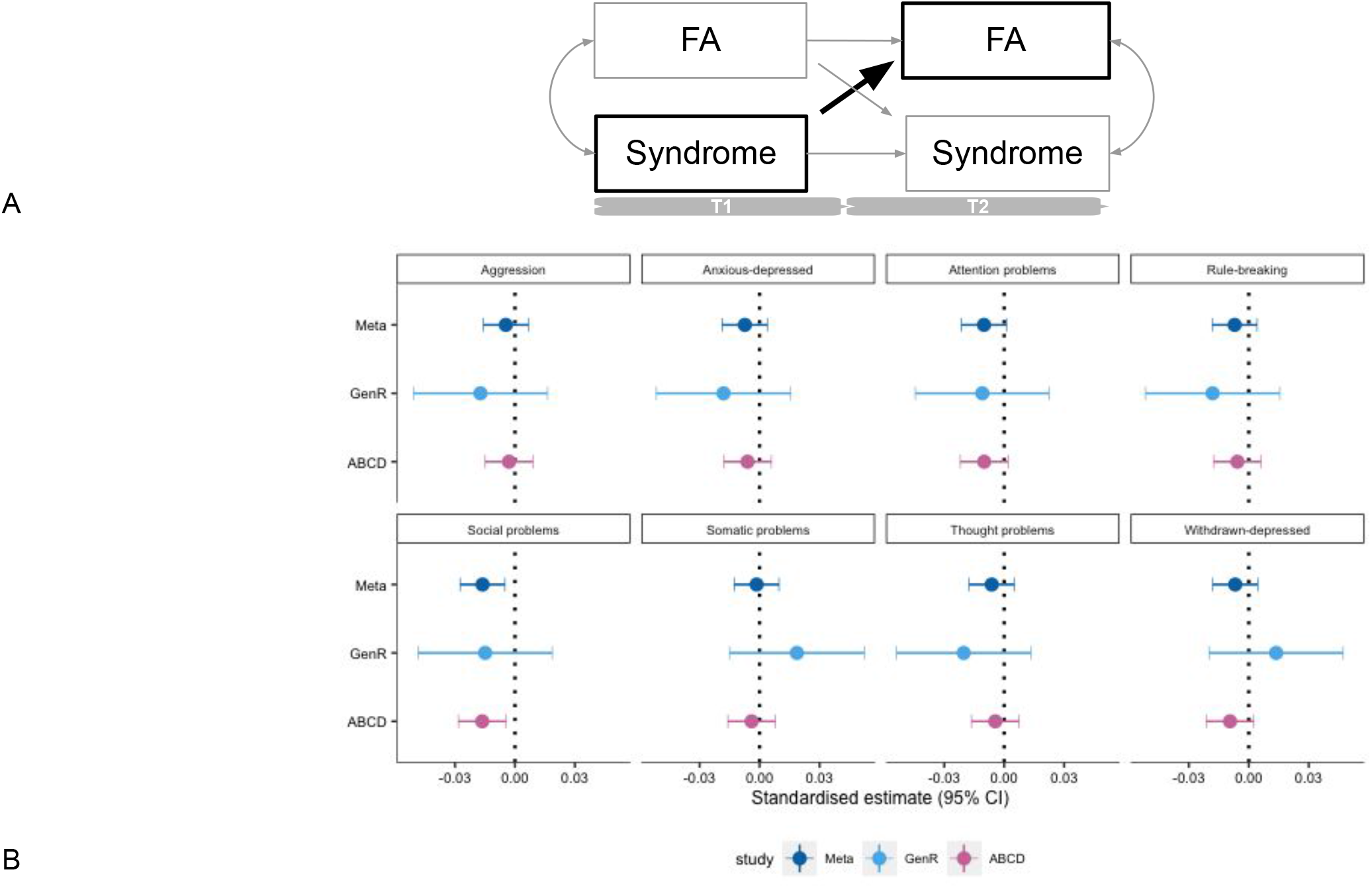
Meta-analysis and single study results for the relation between baseline psychiatric symptoms (syndrome scales) and changes in global FA. *Note*. ABCD = The ABCD Study; GenR = The Generation R Study; FA = fractional anisotropy; Meta = meta-analysis; T = time-point. **A**. Cross-lagged panel model with lagged paths of interest in bold and larger font: psychiatric symptoms T1 → global FA T2 **B**. Standardized coefficients for the effect of syndrome scales at T1 on changes in global FA from T1 to T2. Results are grouped by syndrome scale. Coefficients from the meta-analysis and for each cohort are displayed. Positive coefficients indicate higher psychiatric symptoms at baseline are related to larger changes in FA. Negative coefficients point to higher psychiatric symptoms at baseline being related to lower changes in global FA

Similar results were found in exploratory analyses for *total psychiatric problems* with global FA and MD (**Supplementary Table 8**). No statistically significant lagged path was identified in either temporal direction, effect sizes were modest, and standard errors relatively large (median absolute effect size = 0.007; median SE = 0.008).

We also tested *specific WM tracts* with internalizing and externalizing problems (**Supplementary Table 9**). For brain to behavior, higher FA of the anterior thalamic radiation at baseline predicted larger changes in internalizing and externalizing problems over time (Internalizing: estimate = 0.024, SE = 0.010, *p* = 0.021; Externalizing: estimate = 0.021, SE = 0.010, *p* = 0.037), but did not survive strict FDR correction. For behavior to brain, higher externalizing problems predicted smaller changes in the FA of the corticospinal tract (estimate = -0.013, SE = 0.006, *p* = 0.031), but also did not survive multiple testing correction. Overall, of the 80 tested lagged paths, none survived multiple testing correction (*p*FDR < 0.05) in the meta-analysis. The median absolute effect size was 0.005, with a median standard error of 0.009.

Third, given this lack of evidence for longitudinal associations, we explored whether *cross-sectional* associations of WM microstructure with psychiatric problems were present by observing the covariances at baseline for all tested CLPMs. Some cross-sectional associations were identified and survived multiple testing correction for ABCD: social (estimate = -0.052, SE = 0.015, *p* = 0.0005, *p*FDR = 0.007), thought (estimate = -0.039, SE = 0.015, *p* = 0.009, *p*FDR = 0.043) and attention problems (estimate = -0.041, SE = 0.0150, *p* = 0.007, *p*FDR = 0.043) with global FA, and somatic complaints (estimate = -0.038, SE = 0.015, *p* = 0.011, *p*FDR = 0.043) with global MD. No associations survived multiple testing correction in GenR, but nominal associations were observed for social problems with FA too (estimate = -0.064, SE = 0.030, *p* = 0.035).

Last, we explored whether relations from the main models differed across sexes (**Supplementary Table 10**). No sex differences were found in the structural regression coefficients for ABCD. For GenR, coefficients were different across sexes for internalizing and externalizing problems with FA. Girls and boys showed different directions of effect in such associations: positive for girls and negative for boys. Associations for internalizing problems did not reach statistical significance, but an effect for externalizing problems at baseline on changes in FA in boys was observed (estimate = -0.072, SE = 0.031, *p* = 0.022).

## DISCUSSION

In this large study on WM microstructure and psychiatric symptoms in children and adolescents from the general population, we did not identify any longitudinal relation between internalizing and externalizing problems with global FA and MD in either temporal direction of association (confirmatory analyses). Similarly, no longitudinal associations were observed for total or specific psychiatric problems with global WM microstructure, nor for internalizing and externalizing problems with specific tracts, in either temporal direction, after multiple testing correction (exploratory analyses). However, a few associations were found for specific tracts with internalizing and externalizing problems, and social problems with global WM across cohorts, although only at a nominal significance level. Altogether, we observed small effects, characterized by relatively large standard errors (median absolute estimates: 0.006; median SE: 0.009). Our findings suggest no or very modest longitudinal associations between WM microstructure and psychiatric symptoms from late childhood into early adolescence in the general population, and thus the measures used in this study (e.g., global WM microstructure, broad psychiatric symptom constructs) do not provide additional support for the probabilistic epigenesis assumption of the interactive specialization framework.

Our findings partly contrast prior literature^3,16^. We propose several explanations for our results. First, *differences in design* might have determined distinct results. Generally, studies identifying associations between internalizing/externalizing problems with WM microstructure have used cross-sectional designs. Here we adopted a longitudinal design, which examined how WM associates with changes in psychiatric symptoms over time, accounting for baseline levels of the outcome. This can help dissect effects due to relations of the brain and behavior at earlier ages from relations occurring at a given time-point.

Another reason for our findings might be the chosen *developmental timing*: the transition from late childhood to early adolescence. While adolescence is often considered a key developmental period for psychiatric and brain development^27,42^, there could be downsides in exploring this time window. A large part of brain developmental changes might have already occurred. For instance, grey matter volume peaks before the age of puberty^43^. However, increases in FA and decreases in MD occur well into early adulthood^9^, meaning that investigations in this time period are still likely informative. Moreover, this developmental timing is characterized by pronounced *individual differences*^44,45^. Although with inconsistencies^46^, higher interindividual variability was found from childhood to adulthood^47^. This might explain why longitudinal associations for WM and psychiatric symptoms were found in GenR from early to late childhood^26^, but not from late childhood to early adolescence (this study). The stronger interindividual differences across development might have complicated the identification of associations in adolescence. While we attempted to model individual trajectories with linear mixed-effects models with different starting levels (random intercepts), more repeated measurements are needed to include different rates of development (random slopes). Additionally, it has been suggested that children with certain psychiatric problems present a delayed neurodevelopmental trajectory^11–13^, but that around puberty, there could be reverting to typical neurodevelopment or a shift in associations^11,12^, meaning that relations might not be identifiable at this time-point.

Moreover, the *magnitude of effect sizes* observed here and in previous studies warrants discussion. Median effects of 0.122 were expected for psychiatric problems and WM connectivity in childhood and adolescence, based on prior literature (see https://osf.io/pny92 for power analyses). However, we observed a median absolute effect of 0.009 for our confirmatory analyses and similarly small effect sizes in our exploratory analyses (**Supplementary Figure 2**). Estimates were, therefore, more than an order of magnitude smaller. This is in line with a prior study in ABCD showing lower estimates than expected between commonly established associations such as physical activity and weight (*r* = 0.03)^48^. This might stem from, among other reasons (e.g., differences in samples), the prior wide use of small samples. Small-sized studies are subject to higher sampling variability, potentially determining inflated effect sizes which are more likely to be statistically significant^18^. Compounded by publication bias^49^, the literature may be inadvertently characterized by overly large effects for associations between WM and psychiatric problems across development. With this study, we suggest that such relations from childhood to adolescence in the general population may be smaller than previously expected.

However, a few effect sizes stood out from our exploratory analyses (**Supplementary Figure 2**) and were identified as nominally significant across cohorts: *(i)* the FA of the anterior thalamic radiation in late childhood predicted changes in internalizing and externalizing problems, *(ii)* social problems at baseline predicted changes in global FA, *(iii)* externalizing problems predicted changes in the FA of the corticospinal tract. Notably, these did not survive multiple testing correction and should therefore be carefully evaluated. Yet, multiple testing correction can be overly stringent, determining false-negative results and highlighting the most inflated effects^50^. Replication has thus been suggested as another complementary way to evaluate patterns of results^50^. Our meta-analysis findings may, therefore, be of use for future confirmatory studies. Of note, these associations presented the strongest effects in the meta-analysis. Regardless of our large sample size, since the “true” magnitude of the associations between WM connectivity and psychiatric symptoms remains unknown, our strongest effects may still be inflated. This additionally underscores the importance of using multiple independent large samples.

Further exploring these associations and their temporality remains fundamental to understanding the neurobiological underpinnings and consequences of psychiatric problems. These could advance our knowledge of neurobiological influences on mental health problems and offer key insights to design preventative interventions before deviations in neurodevelopment occur, potentially preventing a cascade of other biological changes. Notably, our findings do not preclude associations being present at different ages (e.g., early to late childhood^26^), in more fine-grained measures of WM (e.g., voxel-based approaches, more advanced diffusion imaging techniques), and at different modalities which might better capture the dynamicity of childhood and adolescence (e.g., functional MRI). We, therefore, highly encourage more investigations into this topic.

Overall, our findings should be evaluated in the context of the considerable strengths of this study. First, we leveraged the *largest samples* to date on neurodevelopment in the general population. Compared to previous studies on the topic, we had, at minimum, a five-fold increase in sample size, which allows for greater power to detect effects as large or even smaller than previously identified. Second, we used *two independent samples* to evaluate the replicability and generalisability of the findings. Third, by adopting *several analytical approaches*, we ensured that our findings were validated and independent of analytical choices. Moreover, with the use of a *dimensional approach* in measuring psychiatric problems, we could examine associations across the entire continuum of psychiatric symptoms.

Several limitations of this study should also be considered. The 2-4 years intervals between assessments could have been insufficient for WM connectivity to influence behavioral trajectories, and vice-versa. Moreover, ABCD and GenR data were not harmonized completely; they have different diffusion MRI pre-processing and quality control pipelines. Additionally, being our samples population-based, only 5-7% of children had severe psychiatric symptoms, based on clinical cut-offs for the CBCL (93^rd^ percentile). This may have diluted associations. Last, our samples are from Western countries in the Global North and include a predominantly highly educated population. Generalizability to other populations (e.g., clinical, Global South, Eastern) should be tested.

In conclusion, we examined longitudinal associations between WM microstructure and psychiatric symptoms by leveraging 5,700 children and adolescents from the general population in the U.S. and Netherlands. While we did not identify any evident bidirectional longitudinal associations between global and specific measures of psychiatric problems and WM connectivity, we offered replicable results, validated with multiple analytical approaches, in two large cohort studies. Our findings might have several implications. First, they might point to the importance of adopting longitudinal designs with repeated measurements and controlling for baseline levels to better dissect which developmental timings might be of key relevance for associations. Second, they might reflect the need for future research to tackle the pubertal period with approaches capturing intraindividual variation across multiple waves of brain and psychiatric assessments. Third, they could highlight the usefulness of leveraging large samples and of replication to minimize the chance of false-positive findings. Further investigations of brain connectivity and psychiatric problems, which take into account the potential for bidirectionality, should be conducted.

## Data Availability

Data for the Generation R Study is available upon request to Prof. Vincent Jaddoe. In line with the European Data Protection Regulation, The Generation R Study data is protected under the General Data Protection Regulation (GDPR). Data for The ABCD Study is already open and available in the NIMH Data Archive (NDA) (nda.nih.gov) to eligible researchers within NIH-verified institutions. Data can be accessed following a data request to the NIH data access committee (https://nda.nih.gov/), which should include information on the planned topic of study. The request is valid for one year. Data use should be in line with the NDA Data Use Certification.

## ACKNOWLEDGEMENTS

As based on the CRediT role taxonomy (https://casrai.org/credit/): conceptualisation (RLM, HT, LDA), data curation (LDA, ABCD and GenR teams (including RLM)), formal Analysis (LDA), funding acquisition (HT, RLM), investigation (LDA), methodology (JvdE, LDA, HT, RLM, Antonio Schettino (AS)), project administration (RLM, HT, LDA), resources (RLM), software (LDA, BX), supervision (RLM, HT), validation (BX), visualization (LDA), writing – original draft (LDA), writing – review & editing (LDA, RLM, HT, BX, JvdE, AS). Statistical expertise for this project was provided by Dr. JvDE. We also thank AS for his support with the open research practices used in this manuscript and for his feedback on the preregistration and manuscript. Additional thanks to Elisabet Blok and Serena Defina for their help with questions on the Generation R and ABCD Studies, and the use of Git, respectively.

## FUNDING

Data used in the preparation of this article were obtained from the Adolescent Brain Cognitive Development^SM^ (ABCD) Study (https://abcdstudy.org), held in the NIMH Data Archive (NDA). This is a multisite, longitudinal study designed to recruit more than 10,000 children age 9-10 and follow them over 10 years into early adulthood. The ABCD Study® is supported by the National Institutes of Health and additional federal partners under award numbers U01DA041048, U01DA050989, U01DA051016, U01DA041022, U01DA051018, U01DA051037, U01DA050987, U01DA041174, U01DA041106, U01DA041117, U01DA041028, U01DA041134, U01DA050988, U01DA051039, U01DA041156, U01DA041025, U01DA041120, U01DA051038, U01DA041148, U01DA041093, U01DA041089, U24DA041123, U24DA041147. A full list of supporters is available at https://abcdstudy.org/federal-partners.html. A listing of participating sites and a complete listing of the study investigators can be found at https://abcdstudy.org/consortium_members/. ABCD consortium investigators designed and implemented the study and/or provided data but did not necessarily participate in the analysis or writing of this report. This manuscript reflects the views of the authors and may not reflect the opinions or views of the NIH or ABCD consortium investigators. The Generation R Study is supported by Erasmus MC, Erasmus University Rotterdam, the Rotterdam Homecare Foundation, the Municipal Health Service Rotterdam area, the Stichting Trombosedienst & Artsenlaboratorium Rijnmond, the Netherlands Organization for Health Research and Development (ZonMw), and the Ministry of Health, Welfare and Sport. Neuroimaging informatics and image analysis was supported by the Sophia Foundation (S18-20, RLM). Netherlands Organization for Scientific Research (Exacte Wetenschappen) and SURFsara (Cartesius Compute Cluster, www.surfsara.nl) supported the Supercomputing resources. LDA, BX, and HT were supported by an NWO-VICI grant (NWO-ZonMW: 016.VICI.170.200 to HT) while RLM was supported by the Sophia Foundation S18-20 and Erasmus MC Fellowship. We thank the participants, general practitioners, hospitals, midwives, and pharmacies in Rotterdam who contributed to the study.

## Notes

### Competing Interest Statement

The authors have declared no competing interest.

### Funding Statement

Data used in the preparation of this article were obtained from the Adolescent Brain Cognitive Development(ABCD)Study (https://abcdstudy.org), held in the NIMH Data Archive (NDA). This is a multisite, longitudinal study designed to recruit more than 10,000 children aged 9-10 and follow them over 10 years into early adulthood. The ABCD Study is supported by the National Institutes of Health and additional federal partners under award numbers U01DA041048, U01DA050989, U01DA051016, U01DA041022, U01DA051018, U01DA051037, U01DA050987, U01DA041174, U01DA041106, U01DA041117, U01DA041028, U01DA041134, U01DA050988, U01DA051039, U01DA041156, U01DA041025, U01DA041120, U01DA051038, U01DA041148, U01DA041093, U01DA041089, U24DA041123, U24DA041147. A full list of supporters is available at https://abcdstudy.org/federal-partners.html. A listing of participating sites and a complete listing of the study investigators can be found at https://abcdstudy.org/consortium_members/. ABCD consortium investigators designed and implemented the study and/or provided data but did not necessarily participate in the analysis or writing of this report. This manuscript reflects the views of the authors and may not reflect the opinions or views of the NIH or ABCD consortium investigators. The Generation R Study is supported by Erasmus MC, Erasmus University Rotterdam, the Rotterdam Homecare Foundation, the Municipal Health Service Rotterdam area, the Stichting Trombosedienst & Artsenlaboratorium Rijnmond, the Netherlands Organization for Health Research and Development (ZonMw), and the Ministry of Health, Welfare and Sport. Neuroimaging informatics and image analysis was supported by the Sophia Foundation (S18-20, RLM). Netherlands Organization for Scientific Research (Exacte Wetenschappen) and SURFsara (Cartesius Compute Cluster, www.surfsara.nl) supported the Supercomputing resources. LDA, BX, and HT were supported by an NWO-VICI grant (NWO-ZonMW: 016.VICI.170.200 to HT) while RLM was supported by the Sophia Foundation S18-20 and Erasmus MC Fellowship.

### Author Declarations

Ethical approval was received from the institutional review boards of the University of California (San Diego) and of each of the 21 data collection sites for ABCD, and the Medical Ethics Committee of Erasmus MC, University Medical Centre (Rotterdam) for GenR. Informed consent or assent has been received from the included participants.

