## Supplementary Materials for "Longitudinal Associations Between White Matter Microstructure and Psychiatric Symptoms in Adolescence"

### SUPPLEMENTARY TEXT

#### METHODS

##### Participants (The ABCD Study)

Participants from the ABCD Study were included/excluded based on the recommended cohort guidelines (imgincl_dmri_include = 1), which involve, among others, no serious MR findings, passed raw and postprocessing quality control, had a minimum of 103 repetitions, and other prespecified cut-offs (see https://osf.io/bg5wt and ABCD Annual Curated Data Release 4.0). Additionally, we excluded participants with clinically relevant incidental findings, similarly to GenR. Two additional exclusion criteria were implemented post-hoc, due to encountered issues, and are described below.

Participants who changed study site from T1 to T2 were excluded (n = 53) due to statistical issues. In fact, we could not include study site information that differed between T1 and T2 due to the high multicollinearity between the sites at T1 and at T2. Therefore, we included effects of site at T1 only in the model (see **Supplementary Table 2** for deviations from preregistered analyses). To minimize the residuals in the estimation of the model-implied matrix vs observed matrix, we excluded children who changed site.

One participant was excluded due to invalid data in the DTI measures (**Supplementary Table 2**). FA values range from 0 to 1. In our samples, the average FA was of 0.50. The excluded child had FA values nearing 1 at T1 (e.g., parahippocampal cingulum $FA_{T1}$ = 1.00, global $FA_{T1}$ = 0.98). While these are in the range of FA, it is highly unlikely to observe such extreme values even in connectivity tracts that are characterised by high directionality of water diffusion. Additionally indicating that the FA values of this child were likely invalid at T1 is a sharp decrease in the child's FA at T2 (e.g., parahippocampal cingulum $FA_{T2}$ = 0.27, global $FA_{T2}$ = 0.46), while children generally show slight increases in the examined time-window.

##### Measures

*Image acquisition*

In ABCD, the high angular resolution diffusion imaging scan was used to obtain diffusion measures. Scanning parameters were harmonised across sites (acquisition matrix = 140 x 140, number of slices = 81, 240 x 240 POV, 100% POV plane, 1.7 x 1.7 x 1.7 mm resolution, 4100 TR, 88 TE, a flip angle of 90°, multiband acceleration = 3, phase partial Fourier = 6/8, 96 diffusion directions, 07:31 acquisition time) and included multiple b-values (500 – 6 dirs., 1000 – 15 dirs., 2000 – 15 dirs., 3000 – 60 dirs.) and a fast integrated $B_0$ distortion correction, implemented with the reversed polarity gradient method[1,2]. In GenR, the DTI data were acquired with an axial spin echo, by using an echo-planar imaging sequence with three b = 0 volumes and 35 diffusion volumes (TR = 12,500 ms, TE = 72.8 ms, FOV = 240 mm x 240 mm, acquisition matrix = 120 x 120, slice thickness = 2mm, voxel size = 2 mm x 2 mm x 2 mm, number of slices = 65, asset acceleration factor = 2, b = 900 s/mm$^2$).

*Image Quality Control*

In ABCD, QC was performed both at the pre-processing and post-processing stages by the ABCD data curation team. During the pre-processing stage, all images were manually reviewed, with images failing QC not undergoing processing. QC at the post-processing stage was done manually in a random sample of 10% of the scans. The obtained scores were used to train Bayesian classifiers which could predict the QC scores of other images based on automated QC metrics. Data considered by the classifier as of low quality underwent additional manual checks (approximately another 10% of images). Outliers were identified by the SPSS Anomaly index procedure. This involves the creation of a clustering model finding natural grouping within a dataset. The models were then used on each image to classify it within a cluster group. Indices of unusualness of the case based on its cluster group were then created. Images were identified as outliers if they were in the top proportion of the sorted anomaly indices.

In GenR, manual and automatic QC was performed for DTI images. In terms of the manual QC[3], visual inspection was conducted and included the examination of sum-of-squares error of the tensor calculation, tract reconstructions, and inter-subject registration accuracy. If a "severe problem" coding was present for either of these measures (e.g., large artifact, motion, problematic registration), the images were considered of low quality and excluded from analyses. Two trained raters assessed each image. For each disagreement, an expert rater was consulted. The eddy tool qc metrics were further used for exclusion. Following previous literature, we adopted a threshold of 3mm of absolute motion[4].

**Covariates**

*The ABCD Study*

All covariates are described in-depth in the ABCD data dictionary (https://nda.nih.gov/data_dictionary.html?source=ABCD%2BRelease%2B3.0&submission=ALL). Age (in months), sex (male, female, not reported), race/ethnicity (White, Black, Hispanic, Asian, other) and parental education were measured in the ABCD Longitudinal Parent Demographics Survey, completed by the primary caregiver. Age was recoded into years. Parental education was initially coded by the ABCD team in 21 categories, ranging from 1st grade to doctoral degree, and was recoded here into three levels (low: 1st to 12th grade, intermediate: high school/GED/college, high: Bachelor's degree or above). Perceived pubertal stage was assessed with child reports on the ABCD Youth Pubertal Development Scale and Menstrual Cycle Survey History and was coded by the ABCD team into pre-, early-, mid-, late-, and post-puberty.

*The Generation R Study*

Child sex (assigned at birth) and age were obtained based on medical records. Child national origin was assessed according to the parent's birth country in a demographic questionnaire based on 11 categories and recoded into 5 major ethnicity categories in the Netherlands (Dutch, Surinamese/Antillean, Turkish/Moroccan, European origin, Other). To assess socioeconomic status, the highest maternal education was used. Maternal education was collected into 6 categories and recoded into 2 (low/intermediate, high) due to the low number of mothers in each category.

Perceived pubertal stage was self-reported by the child at age 14, based on the Pubertal Development Scale questionnaire[5], which includes 8 items (3 for both sexes and an additional 2 for sex-specific pubertal changes) with information on pubertal-related changes such as body hair growth, voice changes, and menstruation. The items are scored from 0 to 4 (e.g., 0 = I do not know, 1 = has not yet started changing, 2 = has barely started changing, 3 = changes are definitely underway, 4 = changes seem complete). Total scores for perceived pubertal development were obtained by following previous literature[5] with minor adjustments. Previous literature suggests averaging the relevant item point scores, without allowing for any missing item, and counting presence of menstruation as 4 points and lack as 1 point. Here, to maximize the number of children with data on pubertal status, we allowed for data on 1 item to be missing (unless such item was the menstruation item due to its high weight on the total score). Additionally, we did not use point values but continuous scores as some children from GenR rated themselves as in between categories for certain items.

#### Statistical Analyses

*Confirmatory analyses: Cross-lagged panel models*

Four cross-lagged panel models were run for each cohort: one for internalising problems with global FA, one for internalising problems with global MD, one for externalising problems with global FA, one for externalising problems with global MD. Therefore, eight lagged coefficients were of interest in each cohort. Both WM microstructure and psychiatric measures were examined as observed variables.

Of note, before data analysis, when necessary, DTI values were scaled to be around the same order of magnitude as the other variables, as is common practice in *lavaan* if some variable variances are much larger or smaller than others. Models included several covariates: sex, ethnicity, parental education, site (for the ABCD Study only), perceived pubertal status, age at MRI (for both time points), and the age difference between the MRI and behavioural assessments at both time points (for GenR only). This was applicable for GenR only as MRI and behavioural assessments were conducted at slightly different child ages, contrarily to the ABCD Study. In line with best practices for running structural equation models, for exogenous (independent) variables, if nominal, they were turned into dummy variables, while ordered factors were transformed into numeric variables (https://lavaan.ugent.be/tutorial/cat.html).

The hypothesised models were compared to competing models, based on likelihood ratio tests. The competing models involved the same model specifications, except for lagged paths being constrained to 0, to test for model equivalence for our paths of interest. Competing models are generally used in structural equation modelling to establish whether the chosen model presents similar model fits to other plausible models[6]. Model re-specifications (e.g., addition/exclusion of a model path) based on modification indices were also considered, where the model fit of the hypothesised models was below pre-specified criteria (RMSEA $\leq$ 0.06; SRMR $\leq$ 0.09) (https://osf.io/pny92). The initially tested cross-lagged panel models did not fit the data sufficiently well. Following modification indices which were in line with previous literature[7], we included an effect of sex on WM microstructure (T1) for GenR. The same model was applied to ABCD, where, additionally, the effect of site on DTI (at T1 and T2) was included. After these modifications, we obtained good model fits in both studies (**Supplementary Table 3**); **Figure 1** displays the final models. To maximise power and highlight replicable results, estimates for each lagged path were standardised and subsequently meta-analysed (fixed-effects, weighted by sample size) using the R package *meta*[8].

*Sensitivity analyses: Linear Mixed Effects Models*

To account for covariate missingness, the *mice* R package for multivariate imputation by chained equations was used[9]. Analyses were run on each dataset, with final estimates for linear mixed-effects models being obtained by pooling the results from all datasets[9]. Linear mixed-effect models with a random intercept were run for ABCD and GenR. Due to the multi-site structure of ABCD, we additionally ran models with site as a main effect or random intercept. Based on likelihood ratio tests, the best fitting model, which included site as a main effect, was selected. Symptoms or WM microstructure over time were regressed on baseline WM microstructure or symptoms respectively, together with selected covariates (age, sex, ethnicity, parental education, puberty), the random intercept, and study-specific variables (site for ABCD and age difference in behavioural and MRI assessment for GenR). An interaction term for time points and symptoms or WM microstructure at baseline was included to investigate our main associations of interest, i.e., the effect of symptoms or WM microstructure on change in WM microstructure and symptoms over time, respectively. This was similar to the cross-lagged panel models and in line with prior literature[3].

*Exploratory analyses*

Eight syndrome scales from the CBCL were tested in association with global FA and MD: attention, thought, and social problems, somatic complaints, and anxious-depressed, withdrawn-depressed, aggressive, and rule-breaking behaviors.

The FA and MD of ten white matter connectivity tracts were tested in association with internalizing/externalizing problems. Such ten connectivity tracts were chosen because they overlapped for ABCD and GenR: *(i)* the cingulate gyrus part of the cingulum, *(ii)* the parahippocampal cingulum, *(iii)* the corticospinal/pyramidal tract, *(iv)* the anterior thalamic radiation, *(v)* the uncinate, *(vi)* the inferior longitudinal fasciculus, *(vii)* the inferior-fronto-occipital fasciculus, *(viii)* the forceps major, *(ix)* the forceps minor, *(x)* the superior longitudinal fasciculus.

To assess whether our results differed across sexes, we reran the main cross-lagged panel models grouped by sex with freely estimated regression coefficients vs. with regression coefficients constrained to equal across sexes. Likelihood ratio tests were used to compare the two models and assess differences across sexes.

**Supplementary Figure 1.** Flowchart for participant inclusion and exclusion for the Generation R and ABCD Studies.

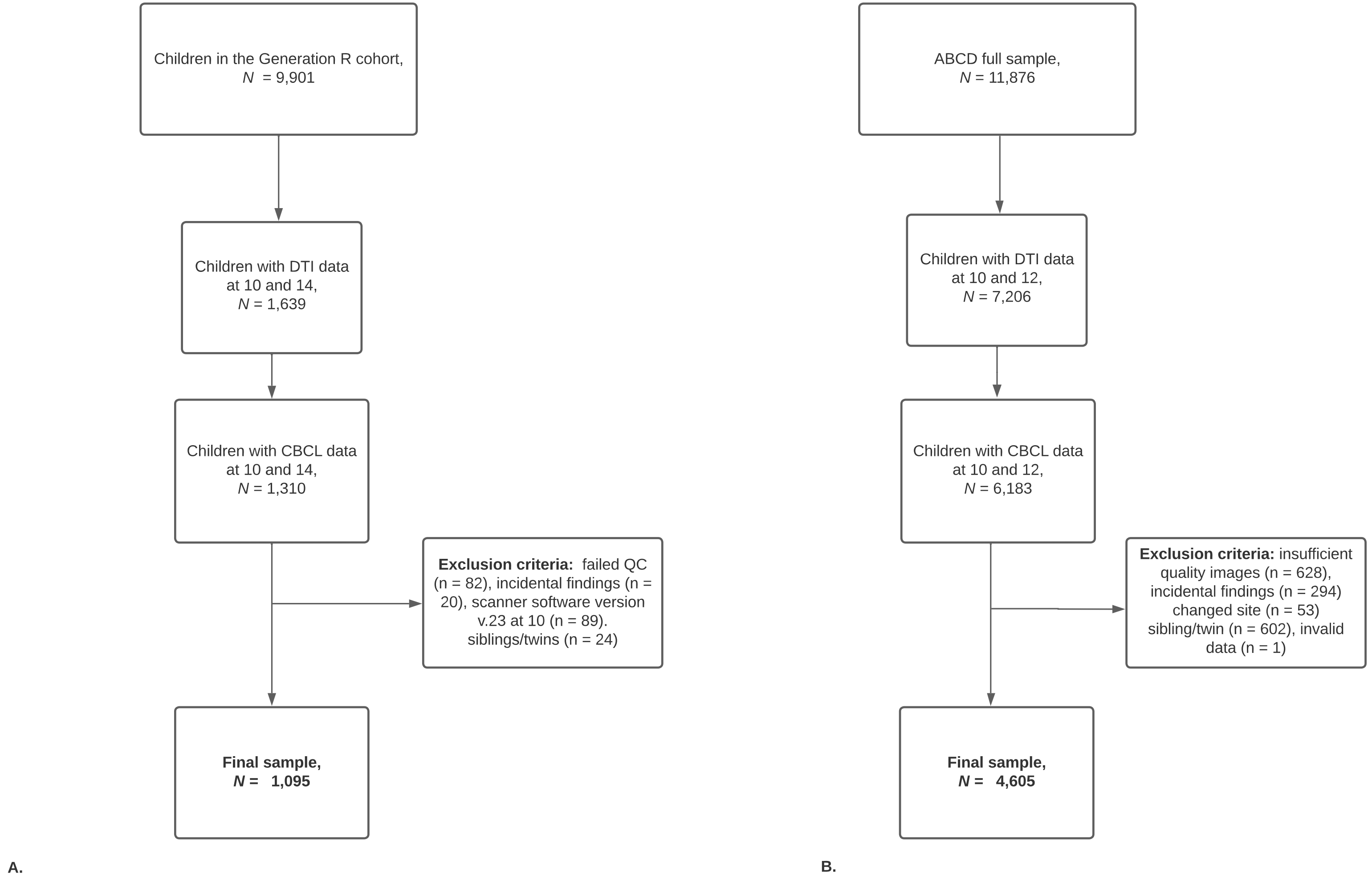


*Note.* **A)** Flowchart for participant inclusion and exclusion for the Generation R Study. **B)** Flowchart for participant inclusion and exclusion for the ABCD Study.

**Supplementary Figure 2.** Histograms of absolute effect sizes (top) and standard errors (bottom) for the exploratory meta-analyses of syndrome scales, tracts, and total problems.

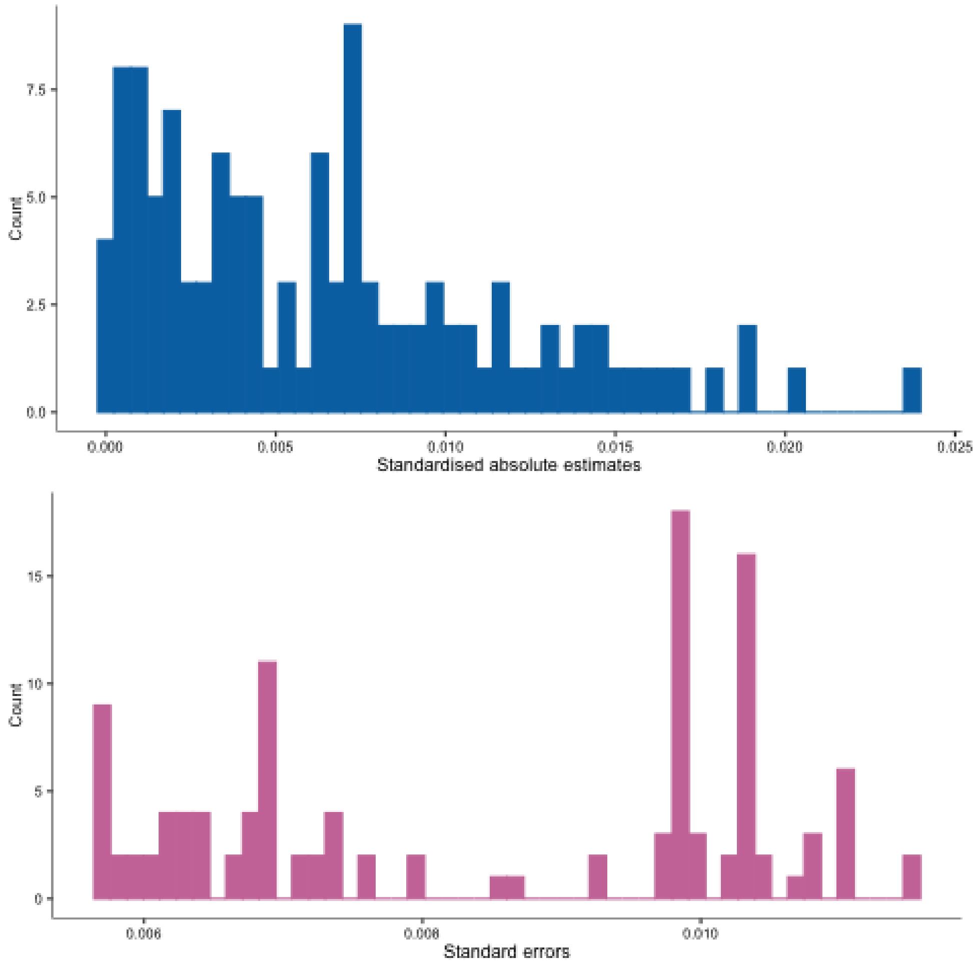

**Supplementary Table 1.** Variables used in this research from the Generation R and ABCD Studies.

| **Dataset / Questionnaire** | **Dataset ID** | **Measures and measures IDs** |
|---|---|---|
| ***The Generation R Study*** | | |
| General data questionnaire | CHILD-ALLGENERALDATA_12112020.sav | Child ID ("idc"), Mother ID ("idm"), siblings / twins ("mother"), ethnicity ("ethninfv3"), sex assigned at birth ("gender"), maternal education ("educm5"), children invited to participate at @9 ("startfase3_9") |
| Child Behavioral checklist at T1 | CHILDCBCL9_incl_Tscores_20201111.sav | <u>Confirmatory analyses:</u><br>Externalising problems ("sum_ext_9m")<br>Internalising problems ("sum_int_9m")<br>Child age at assessment (“agechild_cbcl9m")<br><br><u>Exploratory analyses:</u><br>Anxious depressed ("sum_anx_9m")<br>Withdrawn depressed ("sum_wit_9m")<br>Somatic complaints ("sum_som_9m")<br>Social problems ("sum_sop_9m")<br>Thought problems ("sum_tho_9m")<br>Attention problems ("sum_att_9m")<br>Rule breaking ("sum_rul_9m")<br>Aggression problems ("sum_agg_9m") |
| Child Behavioral checklist at T2 | GR1093-E1_CBCL_18062020.sav | <u>Confirmatory analyses:</u><br>Externalising problems ("sum_ext_14")<br>Internalising problems ("sum_int_14")<br>Child age at assessment ("agechild_gr1093")<br><br><u>Exploratory analyses:</u><br>Anxious depressed ("sum_anx_14")<br>Withdrawn depressed ("sum_wit_14") |

| Dataset / Questionnaire | Dataset ID | Measures and measures IDs |
|---|---|---|
| | | Somatic complaints ("sum_som_14")<br>Social problems ("sum_sop_14")<br>Thought problems ("sum_tho_14")<br>Attention problems ("sum_att_14")<br>Rule breaking ("sum_rul_14")<br>Aggression problems ("sum_agg_14") |
| Pubertal developmental scale questionnaire | GR1095-G_11052020.sav | Puberty items ("g001001r9501_v2", "g002001r9501_v2", "g003001r9501_v2", "g004001r9501_v2", "g005001r9501_v2", "g006001r9501_v2", "g007001r9501_v2") |
| Core MRI data | genr_mri_core_data_20220311.rds | Child age at assessment at @9 (“age_child_mri_f09”)<br>Child age at assessment at @13 ("age_child_mri_f13")<br>Braces at @9 ("has_braces_mri_f09")<br>Braces at @13 ("has_braces_mri_f13")<br>Consent at @9 ("mri_consent_f09")<br>Consent at @13 ("mri_consent_f13")<br>Incidental findings at @9 ("exclude_incidental_f09")<br>Incidental findings at @13 ("exclude_incidental_f13")<br>Software version for at @9 ("mr750_softwareversionshort_dicom")<br>Missingness in global measures at @9 (“missingness_abcd_f09”)<br>Missingness in global measures at @13 (“missingness_abcd_f13”)<br>QC at @9 (“dti_overall_qc_f09”)<br>QC at @13 (“dti_overall_qc_f13”) |
| DTI data | f09_GenR_MRI_eddy_dipy_wls_14Feb2022_autoPtx_dti_stats_inc_glob_measV1.rds | *Global measures:*<br>FA at @9 (“mean_FA_abcd_f09”)<br>MD at @9 (“mean_MD_abcd_f09”)<br>FA at @13 (“mean_FA_abcd_f13”) |

| **Dataset / Questionnaire** | **Dataset ID** | **Measures and measures IDs** |
|---|---|---|
| | f13_GenR_MRI_eddy_dipy_wls_14Feb2022_autoPtx_dti_stats_inc_glob_measV1.rds | MD at @13 (“mean_MD_abcd_f13”)<br>NB measures include the word abcd in them because the mean FA/MD were created based on tracts which were also used in the ABCD Study<br><br>*Tracts right hemisphere FA:*<br><br>Cingulate gyrus (“cgc_r_dti_dipy_wls_wavg_FA”)<br>Parahippocampal cingulum (“cgh_r_dti_dipy_wls_wavg_FA”)<br>Corticospinal tract (“cst_r_dti_dipy_wls_wavg_FA”)<br>Anterior thalamic radiation (“atr_r_dti_dipy_wls_wavg_FA”)<br>Uncinate (“unc_r_dti_dipy_wls_wavg_FA”)<br>Inferior longitudinal fasciculus (“ilf_r_dti_dipy_wls_wavg_FA”)<br>Inferior-fronto-occipital fasciculus (“ifo_r_dti_dipy_wls_wavg_FA”)<br>Superior longitudinal fasciculus (“slf_r_dti_dipy_wls_wavg_FA”)<br><br>*Tracts left hemisphere FA:*<br><br>Cingulate gyrus (“cgc_l_dti_dipy_wls_wavg_FA”)<br>Parahippocampal cingulum (“cgh_l_dti_dipy_wls_wavg_FA”)<br>Corticospinal tract (“cst_l_dti_dipy_wls_wavg_FA”)<br>Anterior thalamic radiation (“atr_l_dti_dipy_wls_wavg_FA”)<br>Uncinate (“unc_l_dti_dipy_wls_wavg_FA”)<br>Inferior longitudinal fasciculus (“ilf_l_dti_dipy_wls_wavg_FA”)<br>Inferior-fronto-occipital fasciculus (“ifo_l_dti_dipy_wls_wavg_FA”)<br>Superior longitudinal fasciculus (“slf_l_dti_dipy_wls_wavg_FA”)<br><br>*Tracts both hemispheres FA:*<br><br>Forceps major (“fma_dti_dipy_wls_wavg_FA”)<br>Forceps minor (“fmi_dti_dipy_wls_wavg_FA”) |

| Dataset / Questionnaire | Dataset ID | Measures and measures IDs |
|---|---|---|
| | | *Tracts right hemisphere MD:*<br><br>Cingulate gyrus (“cgc_r_dti_dipy_wls_wavg_MD”)<br>Parahippocampal cingulum (“cgh_r_dti_dipy_wls_wavg_MD”)<br>Corticospinal tract (“cst_r_dti_dipy_wls_wavg_MD”)<br>Anterior thalamic radiation (“atr_r_dti_dipy_wls_wavg_MD”)<br>Uncinate (“unc_r_dti_dipy_wls_wavg_MD”)<br>Inferior longitudinal fasciculus (“ilf_r_dti_dipy_wls_wavg_MD”)<br>Inferior-fronto-occipital fasciculus (“ifo_r_dti_dipy_wls_wavg_MD”)<br>Superior longitudinal fasciculus (“slf_r_dti_dipy_wls_wavg_MD”)<br><br>*Tracts left hemisphere MD:*<br><br>Cingulate gyrus (“cgc_l_dti_dipy_wls_wavg_MD”)<br>Parahippocampal cingulum (“cgh_l_dti_dipy_wls_wavg_MD”)<br>Corticospinal tract (“cst_l_dti_dipy_wls_wavg_MD”)<br>Anterior thalamic radiation (“atr_l_dti_dipy_wls_wavg_MD”)<br>Uncinate (“unc_l_dti_dipy_wls_wavg_MD”)<br>Inferior longitudinal fasciculus (“ilf_l_dti_dipy_wls_wavg_MD”)<br>Inferior-fronto-occipital fasciculus (“ifo_l_dti_dipy_wls_wavg_MD”)<br>Superior longitudinal fasciculus (“slf_l_dti_dipy_wls_wavg_MD”)<br><br>*Tracts both hemispheres MD:*<br><br>Forceps major (“fma_dti_dipy_wls_wavg_MD”)<br>Forceps minor (“fmi_dti_dipy_wls_wavg_MD”) |
| ***The ABCD Study*** | | |

| Dataset / Questionnaire | Dataset ID | Measures and measures IDs |
|---|---|---|
| ABCD Longitudinal Parent Demographics Survey | abcd_lpds01 | sex (sex), age (interview_age), primary caregiver education (demo_prnt_ed_v2_l), child id (src_subject_id), timepoint (eventname) |
| ABCD ACS Post Stratification Weights | acspsw03 | race/ethnicity (race_ethnicity)<br>Family ID (for exclusion of siblings / twins) (rel_family_id) |
| ABCD Sum Scores Physical Health Youth | abcd_ssphy01 | perceived pubertal status (based on sex at birth) rated by the child (pds_y_ss_female_category_2, pds_y_ss_male_cat_2) |
| ABCD dMRI DTI Full Part 1 | abcd_dmdtifp101 | <u>Confirmatory analyses:</u><br><br>Average FA within all DTI tract fibers (dmdtifp1_38)<br>Average MD within all DTI tract fibers (dmdtifp1_80)<br><br><u>Exploratory analyses:</u><br><br>*Right hemisphere FA:*<br><br>Average FA within DTI atlas tract right cingulate cingulum (dmdtifp1_3)<br>Average FA within DTI atlas tract right parahippocampal cingulum (dmdtifp1_5)<br>Average FA within DTI atlas tract right corticospinal/pyramidal (dmdtifp1_7)<br>Average FA within DTI atlas tract right anterior thalamic radiations (dmdtifp1_9)<br>Average FA within DTI atlas tract right uncinate (dmdtifp1_11)<br>Average FA within DTI atlas tract right inferior longitudinal fasiculus (dmdtifp1_13) |

| Dataset / Questionnaire | Dataset ID | Measures and measures IDs |
|---|---|---|
| | | Average FA within DTI atlas tract right inferior-fronto-occipital fasiculus (dmdtifp1_15)<br>Average FA within DTI atlas tract right superior longitudinal fasiculus (dmdtifp1_20)<br><br>*Left hemisphere FA:*<br><br>Average FA within DTI atlas tract left cingulate cingulum (dmdtifp1_4)<br>Average FA within DTI atlas tract left parahippocampal cingulum (dmdtifp1_6)<br>Average FA within DTI atlas tract left corticospinal/pyramidal (dmdtifp1_8)<br>Average FA within DTI atlas tract left anterior thalamic radiations (dmdtifp1_10)<br>Average FA within DTI atlas tract left uncinate (dmdtifp1_12)<br>Average FA within DTI atlas tract left inferior longitudinal fasiculus (dmdtifp1_14)<br>Average FA within DTI atlas tract left inferior-fronto-occipital fasiculus (dmdtifp1_16)<br>Average FA within DTI atlas tract left superior longitudinal fasiculus (dmdtifp1_21)<br><br>*Both hemispheres FA:*<br><br>Average FA within DTI atlas tract foreceps major (dmdtifp1_17)<br>Average FA within DTI atlas tract foreceps minor (dmdtifp1_18)<br><br>*Right hemisphere MD:*<br><br>Average MD within DTI atlas tract right cingulate cingulum (dmdtifp1_45) |

| **Dataset / Questionnaire** | **Dataset ID** | **Measures and measures IDs** |
|---|---|---|
| | | Average MD within DTI atlas tract right parahippocampal cingulum (dmdtifp1_47)<br>Average MD within DTI atlas tract right corticospinal/pyramidal (dmdtifp1_49)<br>Average MD within DTI atlas tract right anterior thalamic radiations (dmdtifp1_51)<br>Average MD within DTI atlas tract right uncinate (dmdtifp1_53)<br>Average MD within DTI atlas tract right inferior longitudinal fasiculus (dmdtifp1_55)<br>Average MD within DTI atlas tract right inferior-fronto-occipital fasiculus (dmdtifp1_57)<br>Average MD within DTI atlas tract right superior longitudinal fasiculus (dmdtifp1_62)<br><br>*Both hemispheres MD:*<br><br>Average MD within DTI atlas tract foreceps major (dmdtifp1_59)<br>Average MD within DTI atlas tract foreceps minor (dmdtifp1_60)<br><br>*Left hemisphere MD:*<br><br>Average MD within DTI atlas tract left cingulate cingulum (dmdtifp1_46)<br>Average MD within DTI atlas tract left parahippocampal cingulum (dmdtifp1_48)<br>Average MD within DTI atlas tract left corticospinal/pyramidal (dmdtifp1_50)<br>Average MD within DTI atlas tract left anterior thalamic radiations (dmdtifp1_52)<br>Average MD within DTI atlas tract left uncinate (dmdtifp1_54)<br>Average MD within DTI atlas tract left inferior longitudinal fasiculus (dmdtifp1_56) |

| Dataset / Questionnaire | Dataset ID | Measures and measures IDs |
|---|---|---|
| | | Average MD within DTI atlas tract left inferior-fronto-occipital fasiculus (dmdtifp1_58)<br>Average MD within DTI atlas tract left superior longitudinal fasiculus (dmdtifp1_63) |
| ABCD Parent Child Behavior Checklist Scores Aseba (CBCL) | abcd_cbcls01 | Confirmatory analyses:<br><br>Internalizing raw score (cbcl_scr_syn_internal_r),<br>Externalizing raw score (cbcl_scr_syn_external_r)<br><br>Exploratory analyses:<br><br>Anxious depressed (cbcl_scr_syn_anxdep_r)<br>Withdrawn depressed (cbcl_scr_syn_withdep_r)<br>Somatic complaints (cbcl_scr_syn_somatic_r)<br>Social problems (cbcl_scr_syn_social_r)<br>Thought problems (cbcl_scr_syn_thought_r)<br>Attention problems (cbcl_scr_syn_attention_r)<br>Rule breaking (cbcl_scr_syn_rulebreak_r)<br>Aggressive problems (cbcl_scr_syn_aggressive_r) |
| ABCD Recommended Imaging Inclusion | abcd_imgincl01 | Images inclusion for DTI (imgincl_dmri_include) |
| ABCD Longitudinal tracking | abcd_lt01 | Site id for scanning at each visit (site_id_l) |
| ABCD MR findings | abcd_mrfindings02 | Incidental findings (mrif_score) |

**Supplementary Table 2.** Deviations from the preregistered analyses

| Preregistered analysis | Deviation | Reason for deviation |
|---|---|---|
| *Sample participants or measures* | | |
| Including data just from the baseline wave and from the 2-year follow up wave for the Generation R and ABCD Studies | We included data on one covariate from the 1 year follow up wave for ABCD | Data on parental education was not available at the baseline wave, but just at the 2 year and 1 year follow up waves. We decided to include the parental education data from the 1 year follow up wave as this precedes temporally the 2-year follow up outcomes, which makes confounding bias assumptions about temporality of associations more plausible. |
| Including data on pubertal stage (time point not specified, rater not specified) in the ABCD Study | We realized that data on pubertal stage is present both for baseline and 2-year follow up, rated both by the child and parent. We decided to use the data from the 2-year follow up rated by the child. | Data on pubertal stage is important for the model and generally varies across time due to the child development. For consistency purposes, since the Generation R Study had data just at T2 on puberty, we selected pubertal data at T2 for the ABCD Study too. Additionally, we noticed that there was not one aggregated variable for child and parent rated data, as we initially thought. We therefore decided to use child reports, in line with what was available for the Generation R cohort. |
| Log transformation of the skewed behavioral data | We square-root transformed the behavioral data | This was due to a misunderstanding between co-authors. Square-root, as opposed to log, transformation is generally used for psychiatric problems as rated in the CBCL. The first author was not aware of this while writing the preregistration. |
| Participants in the ABCD Study | Excluded 30 participants who changed site from baseline to the two-year follow up | We initially included site in the CLPM predicting white matter microstructure at baseline (T1), but the modification indices suggested for site to load on white matter microstructure at follow up too (T2). However, we discovered that 30 children changed site from time 1 to time |

| Preregistered analysis | Deviation | Reason for deviation |
|---|---|---|
| | | 2. We did not predict this in our preregistration and decided to exclude these participants to ensure more preciseness. Of note, we could not include site from time 1 and 2 in our analyses because the variables were largely overlapping and thus multicollinearity occurred and the model could not converge. |
| No additional data exclusion in ABCD as the ABCD Data Team already checked for invalid data | Excluded 1 participant with invalid FA data. Such invalid data was not corrected in the ABCD data release 4.0. | FA is a measure of directionality of water diffusion which ranges from 0 to 1. Both at T1 and T2 children had, on average, an FA of 0.50-0.51. Here, we found a child with a global FA nearing 1, likely indicating an invalid entry. See Supplementary Methods, *Participants* for more information |
| Categorisation of maternal education into 3 categories and of ethnicity into 6 for the Generation R Study | 5 categories for ethnicity<br>2 for maternal education | There were very few participants in the low education category, so we collapsed it together with the intermediate education category. Likewise for the Turkish and Moroccan categories. |
| Pubertal developmental scale calculation as sum score in the Generation R Study | This was calculated as an average score weighted by the number of available items | We noticed only when creating the variable that average scores should be created rather than sum scores, based on the protocol of the Pubertal Development Scale[5]. |
| *Confirmatory analyses* | | |
| Include the age difference between MRI and behavioural assessments in the models in both the Generation R and ABCD Studies | Age difference between MRI and behavioural assessments at the same time points were not included in the model for the ABCD Study | Within the ABCD Study, the MRI and behavioural assessments were conducted at the same child age. There was therefore no need to look at the age difference. We were not aware of this at the time of preregistration. |
| Evaluation of the results based on the most parsimonious and best fitting models | Evaluated the results based on the best fitting model version of the hypothesized model | The competing models were more parsimonious and not significantly different in model fit from the hypothesized models. Based on the preregistration, we should have |

| **Preregistered analysis** | **Deviation** | **Reason for deviation** |
|---|---|---|
| | | evaluated our results based on the competing models. However, these did not include lagged paths, which contain the coefficients of interest for our confirmatory analyses. While certainly, this already indicated that the lagged paths were not of high importance in the model and thus that our results were negative, interpreting results based on the competing models would not inform us on the coefficients of the paths of interest. We therefore evaluated the results based on the best fitting model version (i.e., after modification indices) of the hypothesized model. |
| *Sensitivity and exploratory analyses* | | |
| Robustness checks run for significant findings | Ran robustness checks for all results, regardless of their statistical significance | After consultation with an open research expert, we decided to run robustness checks regardless of the results which we obtained. This is because a positive or negative result should be confirmed by multiple methods, to ensure that it is not dependent on specific statistical choices. |
| Running additional adjustments for covariates in the CLPM | We did not re-run CLPMs including additional covariates | Such sensitivity analysis was envisioned to account for potential confounding bias by factors which were not initially considered in the CLPM, as is common epidemiological practice. As our results were negative, we did not deem necessary to consider further covariates. In fact, we did not expect negative confounding (i.e., that novel associations would appear after further covariates were included) |
| Testing 12 tracts for the Generation R and ABCD Studies for our exploratory analyses | We tested 10 tracts which overlapped between the Generation R and ABCD Studies | Because we aimed to meta-analyse the results from the tested tracts, it was necessary to test the tracts which were overlapping between the two studies. We noticed, upon |

| Preregistered analysis | Deviation | Reason for deviation |
|---|---|---|
| | | checking the data, that the Generation R and ABCD studies had overlapping data at 10 tracts only. |

**Supplementary Table 3.** Model fits for the Generation R and ABCD Studies.

| **Model** | **RMSEA** | **SRMR** | **CFI** | **TLI** |
|---|---|---|---|---|
| **The ABCD Study** | | | | |
| Internalising - FA | 0.025 | 0.010 | 0.988 | 0.977 |
| Internalising - MD | 0.024 | 0.010 | 0.986 | 0.975 |
| Externalising - FA | 0.029 | 0.012 | 0.984 | 0.971 |
| Externalising - MD | 0.029 | 0.012 | 0.982 | 0.966 |
| **The Generation R Study** | | | | |
| Internalising - FA | 0.037 | 0.025 | 0.974 | 0.959 |
| Internalising - MD | 0.047 | 0.022 | 0.963 | 0.942 |
| Externalising - FA | 0.044 | 0.026 | 0.963 | 0.942 |
| Externalising - MD | 0.053 | 0.024 | 0.953 | 0.926 |

*Note.* CFI = comparative fit index; FA = fractional anisotropy; MD = mean diffusivity; RMSEA = root mean square error of approximation; SRMR = Standardized Root Mean Square Residual; TLI = Tucker-Lewis index

**Supplementary Table 4.** Results from the linear-mixed effects models for the ABCD and Generation R Studies.

| | | The ABCD Study | | | The Generation R Study | | |
|---|---|---|---|---|---|---|---|
| **Predictor (T1)** | **Outcome (T1, T2)** | **Est** | **SE** | ***P*** | **Est** | **SE** | ***P*** |
| Int | FA | 0.000 | 0.000 | 0.454 | 0.000 | 0.000 | 0.801 |
| | MD | -0.000 | 0.000 | 0.420 | 0.000 | 0.000 | 0.692 |
| Ext | FA | 0.000 | 0.000 | 0.189 | 0.000 | 0.000 | 0.803 |
| | MD | -0.000 | 0.000 | 0.036 | 0.000 | 0.000 | 0.447 |
| FA | Int | 0.124 | 0.557 | 0.824 | 1.181 | 2.268 | 0.603 |
| | Ext | -0.127 | 0.547 | 0.816 | 2.489 | 2.318 | 0.283 |
| MD | Int | 0.847 | 0.804 | 0.292 | -0.468 | 1.806 | 0.796 |
| | Ext | -0.063 | 0.790 | 0.936 | -1.420 | 1.847 | 0.442 |

*Note.* Estimates indicate the relation between baseline levels of the predictor and change of the outcome over time (i.e., the interaction between the predictor and time-point) and are unstandardized. CI = confidence interval; Est = estimate; FA = fractional anisotropy; FDR = false discovery rate; MD = mean diffusivity; *p* = *p*-value; T1 = time point 1 (baseline assessment); T2 = time point 2 (follow up assessment)

**Supplementary Table 5.** Likelihood ratio test for model equivalence between CLPMs with and without lagged paths

| | **Model** | **Df** | **AIC** | **BIC** | $\chi^2$ | $\chi^2$ **diff** | **Df diff** | ***p*** |
|---|---|---|---|---|---|---|---|---|
| **The ABCD Study** | | | | | | | | |
| Internalising - FA | Hypothesized | 86 | 49373 | 52642 | 2906.9 | | | |
| | Competing | 88 | 49370 | 52627 | 2908.5 | 1.562 | 2 | 0.458 |
| Internalising - MD | Hypothesized | 86 | 41917 | 45186 | 1365.0 | | | |
| | Competing | 88 | 41915 | 45171 | 1367.0 | 1.973 | 2 | 0.373 |
| Externalising - FA | Hypothesized | 86 | 49864 | 53133 | 2972.4 | | | |
| | Competing | 88 | 49862 | 53118 | 2974.0 | 1.648 | 2 | 0.439 |
| Externalising - MD | Hypothesized | 86 | 42408 | 45677 | 1438.1 | | | |
| | Competing | 88 | 42405 | 45661 | 1439.6 | 1.460 | 2 | 0.482 |
| **The Generation R Study** | | | | | | | | |
| Internalising - FA | Hypothesized | 33 | 22126 | 22636 | 87.4 | | | |
| | Competing | 35 | 22122 | 22622 | 87.4 | 0.050 | 2 | 0.975 |
| Internalising - MD | Hypothesized | 33 | 22823 | 23332 | 161.5 | | | |
| | Competing | 35 | 22819 | 23319 | 161.6 | 0.120 | 2 | 0.942 |
| Externalising - FA | Hypothesized | 33 | 22187 | 22697 | 107.2 | | | |
| | Competing | 35 | 22184 | 22684 | 108.3 | 1.085 | 2 | 0.581 |
| Externalising - MD | Hypothesized | 33 | 22187 | 22697 | 107.2 | | | |
| | Competing | 35 | 22184 | 22684 | 108.3 | 1.085 | 2 | 0.581 |

*Note.* AIC = Akaike's Information Criteria; BIC = Bayesian Information Criteria; CLPM = cross-lagged panel model; diff = difference; df = degrees of freedom; FA = fractional anisotropy; MD = mean diffusivity; *p* = *p*-value; $\chi^2$ = chi-square

**Supplementary Table 6.** Non-response analysis for the Generation R Study

| **Variable** | ***t* or χ statistic** | **df** | ***p*-value** |
|---|---|---|---|
| Puberty | -0.765 | 1337.762 | 0.444 |
| Sex | 0.140 | 1 | 0.709 |
| Ethnicity | 1.893 | 4 | 0.755 |
| Maternal education | 0.849 | 1 | 0.357 |

*Note.* Df = degrees of freedom; *t* = t-score; χ = chi-square

**Supplementary Table 7.** Results for exploratory analyses for each syndrome scale with global FA and MD

| Model | Meta-analysis | | | | ABCD | | | GenR | | |
|---|---|---|---|---|---|---|---|---|---|---|
| | **Estimate** | **SE** | ***p*** | **FDR** | **Estimate** | **SE** | ***p*** | **Estimate** | **SE** | ***p*** |
| ***Global FA T1 → syndrome scales T2*** | | | | | | | | | | |
| Anxious/Depressed T2 ~ FA T1 | 0.015 | 0.011 | 0.160 | 0.690 | 0.015 | 0.012 | 0.210 | 0.017 | 0.026 | 0.520 |
| Withdrawn/Depressed T2 ~ FA T1 | 0.007 | 0.011 | 0.499 | 0.806 | 0.005 | 0.012 | 0.653 | 0.016 | 0.026 | 0.530 |
| Somatic Complaints T2 ~ FA T1 | -0.001 | 0.012 | 0.954 | 0.966 | 0.004 | 0.013 | 0.739 | -0.023 | 0.027 | 0.395 |
| Social Problems T2 ~ FA T1 | 0.003 | 0.011 | 0.794 | 0.945 | -0.001 | 0.012 | 0.939 | 0.021 | 0.026 | 0.426 |
| Thought Problems T2 ~ FA T1 | 0.014 | 0.011 | 0.194 | 0.690 | 0.011 | 0.012 | 0.377 | 0.031 | 0.026 | 0.236 |
| Attention Problems T2 ~ FA T1 | -0.001 | 0.010 | 0.893 | 0.955 | 0.003 | 0.011 | 0.782 | -0.023 | 0.024 | 0.341 |
| Aggressive Behavior T2 ~ FA T1 | 0.007 | 0.010 | 0.503 | 0.806 | 0.008 | 0.011 | 0.450 | -0.001 | 0.025 | 0.954 |
| Rule-Breaking Behavior T2 ~ FA T1 | 0.006 | 0.011 | 0.580 | 0.867 | 0.003 | 0.012 | 0.834 | 0.025 | 0.027 | 0.370 |
| ***Syndrome scales T1 → Global FA T2*** | | | | | | | | | | |
| FA T2 ~ Anxious/Depressed T1 | -0.007 | 0.006 | 0.199 | 0.690 | -0.006 | 0.006 | 0.323 | -0.018 | 0.017 | 0.293 |
| FA T2 ~ Withdrawn/Depressed T1 | -0.007 | 0.006 | 0.234 | 0.690 | -0.009 | 0.006 | 0.122 | 0.014 | 0.017 | 0.424 |
| FA T2 ~ Somatic Complaints T1 | -0.001 | 0.006 | 0.797 | 0.9405 | -0.004 | 0.006 | 0.512 | 0.019 | 0.017 | 0.278 |
| FA T2 ~ Social Problems T1 | -0.016 | 0.006 | 0.005 | 0.148 | -0.016 | 0.006 | 0.007 | -0.015 | 0.017 | 0.384 |
| FA T2 ~ Thought Problems T1 | -0.006 | 0.006 | 0.278 | 0.690 | -0.004 | 0.006 | 0.465 | -0.020 | 0.017 | 0.237 |
| FA T2 ~ Attention Problems T1 | -0.010 | 0.006 | 0.077 | 0.618 | -0.010 | 0.006 | 0.099 | -0.011 | 0.017 | 0.523 |
| FA T2 ~ Aggressive Behavior T1 | -0.005 | 0.006 | 0.426 | 0.757 | -0.003 | 0.006 | 0.626 | -0.017 | 0.017 | 0.314 |
| FA T2 ~ Rule-Breaking Behavior T1 | -0.007 | 0.006 | 0.216 | 0.690 | -0.006 | 0.006 | 0.349 | -0.018 | 0.017 | 0.291 |
| ***Global MD T1 → Syndrome scales T2*** | | | | | | | | | | |

| | Meta-analysis | | | | ABCD | | | GenR | | |
|---|---|---|---|---|---|---|---|---|---|---|
| Anxious/Depressed T2 ~ MD T1 | 0.010 | 0.011 | 0.367 | 0.739 | 0.012 | 0.012 | 0.309 | -0.002 | 0.026 | 0.948 |
| Withdrawn/Depressed T2 ~ MD T1 | 0.002 | 0.011 | 0.846 | 0.955 | 0.004 | 0.012 | 0.751 | -0.006 | 0.026 | 0.828 |
| Somatic Complaints T2 ~ MD T1 | 0.013 | 0.012 | 0.268 | 0.690 | 0.014 | 0.013 | 0.286 | 0.009 | 0.028 | 0.741 |
| Social Problems T2 ~ MD T1 | 0.015 | 0.011 | 0.154 | 0.690 | 0.020 | 0.012 | 0.086 | -0.009 | 0.026 | 0.734 |
| Thought Problems T2 ~ MD T1 | 0.009 | 0.011 | 0.414 | 0.757 | 0.015 | 0.012 | 0.204 | -0.021 | 0.026 | 0.419 |
| Attention Problems T2 ~ MD T1 | 0.019 | 0.010 | 0.056 | 0.603 | 0.020 | 0.011 | 0.059 | 0.011 | 0.024 | 0.657 |
| Aggressive Behavior T2 ~ MD T1 | 0.009 | 0.010 | 0.369 | 0.739 | 0.011 | 0.011 | 0.305 | -0.003 | 0.025 | 0.910 |
| Rule-Breaking Behavior T2 ~ MD T1 | 0.001 | 0.011 | 0.895 | 0.955 | 0.001 | 0.012 | 0.905 | 0.002 | 0.028 | 0.955 |
| ***Syndrome scales T1 → Global MD T2*** | | | | | | | | | | |
| MD T2 ~ Anxious/Depressed T1 | 0.006 | 0.007 | 0.363 | 0.739 | 0.006 | 0.008 | 0.472 | 0.009 | 0.016 | 0.551 |
| MD T2 ~ Withdrawn/Depressed T1 | 0.002 | 0.007 | 0.772 | 0.945 | 0.002 | 0.008 | 0.819 | 0.003 | 0.016 | 0.848 |
| MD T2 ~ Somatic Complaints T1 | -0.013 | 0.007 | 0.051 | 0.603 | -0.013 | 0.008 | 0.089 | -0.015 | 0.016 | 0.336 |
| MD T2 ~ Social Problems T1 | 0.004 | 0.007 | 0.596 | 0.867 | 0.003 | 0.008 | 0.654 | 0.005 | 0.016 | 0.772 |
| MD T2 ~ Thought Problems T1 | 0.003 | 0.007 | 0.642 | 0.893 | 0.001 | 0.008 | 0.898 | 0.012 | 0.016 | 0.430 |
| MD T2 ~ Attention Problems T1 | 0.002 | 0.007 | 0.755 | 0.945 | 0.003 | 0.008 | 0.711 | -0.001 | 0.016 | 0.963 |
| MD T2 ~ Aggressive Behavior T1 | 0.000 | 0.007 | 0.966 | 0.966 | -0.005 | 0.008 | 0.525 | 0.019 | 0.016 | 0.233 |
| MD T2 ~ Rule-Breaking Behavior T1 | -0.007 | 0.007 | 0.280 | 0.690 | -0.012 | 0.008 | 0.132 | 0.010 | 0.016 | 0.540 |

*Note.* ~ = regressed on; CI = confidence interval; FA = fractional anisotropy; FDR = false discovery rate; MD = mean diffusivity; SE = standard error; T = time-point.

**Supplementary Table 8.** Results for exploratory analyses for global FA and MD with total problems.

| | Meta-analysis | | | | ABCD | | | GenR | | |
|---|---|---|---|---|---|---|---|---|---|---|
| **Model** | **Estimate** | **SE** | ***p*** | **FDR** | **Estimate** | **SE** | ***p*** | **Estimate** | **SE** | ***p*** |
| Total Problems T2 ~ FA T1 | 0.003 | 0.009 | 0.761 | 0.845 | 0.001 | 0.01 | 0.884 | 0.009 | 0.022 | 0.679 |
| Total Problems T2 ~ MD T1 | 0.012 | 0.009 | 0.205 | 0.41 | 0.016 | 0.01 | 0.105 | -0.012 | 0.023 | 0.607 |
| FA T2 ~ Total Problems T1 | -0.01 | 0.006 | 0.069 | 0.276 | -0.01 | 0.006 | 0.097 | -0.013 | 0.017 | 0.445 |
| MD T2~Total Problems T1 | -0.001 | 0.007 | 0.845 | 0.845 | -0.004 | 0.008 | 0.606 | 0.009 | 0.016 | 0.548 |

*Note.* ~ = regressed on; FA = fractional anisotropy; MD = mean diffusivity; *p* = *p*-value; SE = standard error; T = time-point.

**Supplementary Table 9.** Results for exploratory analyses for each white matter connectivity tract with internalising and externalising problems.

| Model | Meta-analysis | | | | ABCD | | | GenR | | |
|---|---|---|---|---|---|---|---|---|---|---|
| | Estimate | SE | *p* | FDR | Estimate | SE | *p* | Estimate | SE | *p* |
| ***Tract-based FA T1 → Psychiatric symptoms T2*** | | | | | | | | | | |
| Internalising T2 ~ Cingulate Gyrus FA T1 | 0.019 | 0.010 | 0.068 | 0.968 | 0.009 | 0.011 | 0.438 | 0.064 | 0.024 | 0.008 |
| Internalising T2 ~ Parahippocampal Cingulum FA T1 | 0.012 | 0.010 | 0.237 | 0.968 | 0.016 | 0.011 | 0.165 | -0.005 | 0.025 | 0.851 |
| Internalising T2 ~ Corticospinal Tract FA T1 | -0.001 | 0.010 | 0.947 | 0.995 | 0.000 | 0.011 | 0.98 | -0.005 | 0.024 | 0.833 |
| Internalising T2 ~ Anterior Thalamic Radiation FA T1 | 0.024 | 0.010 | 0.021 | 0.968 | 0.021 | 0.011 | 0.063 | 0.036 | 0.024 | 0.138 |
| Internalising T2 ~ Uncinate FA T1 | 0.007 | 0.010 | 0.492 | 0.995 | 0.010 | 0.011 | 0.378 | -0.007 | 0.024 | 0.790 |
| Internalising T2 ~ Inferior Longitudinal Fasciculus FA T1 | 0.012 | 0.010 | 0.263 | 0.968 | 0.012 | 0.011 | 0.299 | 0.011 | 0.025 | 0.673 |
| Internalising T2 ~ Inferior-fronto-occipital Fasciculus FA T1 | 0.011 | 0.010 | 0.292 | 0.968 | 0.012 | 0.011 | 0.281 | 0.004 | 0.025 | 0.859 |
| Internalising T2 ~ Superior Longitudinal Fasciculus FA T1 | 0.001 | 0.010 | 0.940 | 0.995 | 0.011 | 0.011 | 0.347 | -0.045 | 0.025 | 0.065 |
| Internalising T2 ~ Forceps Major FA T1 | 0.001 | 0.010 | 0.911 | 0.995 | 0.002 | 0.011 | 0.836 | -0.004 | 0.025 | 0.855 |
| Internalising T2 ~ Forceps Minor FA T1 | 0.000 | 0.010 | 0.971 | 0.995 | -0.001 | 0.011 | 0.936 | 0.006 | 0.024 | 0.795 |
| Externalising T2 ~ Cingulate Gyrus FA T1 | 0.011 | 0.010 | 0.272 | 0.968 | 0.007 | 0.011 | 0.497 | 0.029 | 0.025 | 0.234 |
| Externalising T2 ~ Parahippocampal Cingulum FA T1 | 0.004 | 0.010 | 0.700 | 0.995 | 0.009 | 0.011 | 0.391 | -0.026 | 0.025 | 0.307 |
| Externalising T2 ~ Corticospinal Tract FA T1 | 0.001 | 0.010 | 0.957 | 0.995 | -0.005 | 0.011 | 0.661 | 0.028 | 0.025 | 0.252 |
| Externalising T2 ~ Anterior Thalamic Radiation FA T1 | 0.021 | 0.010 | 0.037 | 0.968 | 0.020 | 0.011 | 0.063 | 0.024 | 0.025 | 0.332 |
| Externalising T2 ~ Uncinate FA T1 | 0.010 | 0.010 | 0.312 | 0.968 | 0.011 | 0.011 | 0.286 | 0.002 | 0.025 | 0.932 |
| Externalising T2 ~ Inferior Longitudinal Fasciculus FA T1 | 0.007 | 0.010 | 0.448 | 0.995 | 0.012 | 0.011 | 0.270 | -0.017 | 0.025 | 0.509 |
| Externalising T2 ~ Inferior-fronto-occipital Fasciculus FA T1 | -0.003 | 0.010 | 0.731 | 0.995 | -0.001 | 0.011 | 0.904 | -0.015 | 0.025 | 0.557 |
| Externalising T2 ~ Superior Longitudinal Fasciculus FA T1 | 0.004 | 0.010 | 0.712 | 0.995 | 0.006 | 0.011 | 0.547 | -0.011 | 0.025 | 0.645 |

| | Meta-analysis | | | | ABCD | | | GenR | | |
|---|---|---|---|---|---|---|---|---|---|---|
| Externalising T2 ~ Forceps Major FA T1 | -0.002 | 0.010 | 0.858 | 0.995 | -0.002 | 0.011 | 0.836 | 0.001 | 0.025 | 0.976 |
| Externalising T2 ~ Forceps Minor FA T1 | -0.001 | 0.010 | 0.909 | 0.995 | 0.000 | 0.011 | 1.000 | -0.007 | 0.025 | 0.773 |
| ***Psychiatric symptoms T1 → Tract-based FA T2*** | | | | | | | | | | |
| Cingulate Gyrus FA T2 ~ Internalising T1 | -0.004 | 0.007 | 0.520 | 0.995 | -0.008 | 0.007 | 0.283 | 0.023 | 0.020 | 0.261 |
| Parahippocampal Cingulum FA T2 ~ Internalising T1 | -0.003 | 0.006 | 0.574 | 0.995 | -0.001 | 0.006 | 0.856 | -0.027 | 0.020 | 0.181 |
| Corticospinal Tract FA T2 ~ Internalising T1 | -0.003 | 0.006 | 0.584 | 0.995 | -0.007 | 0.006 | 0.272 | 0.028 | 0.019 | 0.127 |
| Anterior Thalamic Radiation FA T2 ~ Internalising T1 | -0.001 | 0.006 | 0.895 | 0.995 | 0.001 | 0.007 | 0.910 | -0.015 | 0.020 | 0.457 |
| Uncinate FA T2 ~ Internalising T1 | 0.000 | 0.006 | 0.982 | 0.995 | 0.000 | 0.007 | 0.957 | -0.001 | 0.018 | 0.94 |
| Inferior Longitudinal Fasciculus FA T2 ~ Internalising T1 | -0.008 | 0.006 | 0.230 | 0.968 | -0.009 | 0.007 | 0.160 | 0.005 | 0.017 | 0.777 |
| Inferior-fronto-occipital Fasciculus FA T2 ~ Internalising T1 | 0.000 | 0.006 | 0.974 | 0.995 | -0.004 | 0.006 | 0.573 | 0.024 | 0.016 | 0.129 |
| Superior Longitudinal Fasciculus FA T2 ~ Internalising T1 | -0.006 | 0.006 | 0.349 | 0.968 | -0.007 | 0.007 | 0.266 | 0.003 | 0.015 | 0.846 |
| Forceps Major FA T2 ~ Internalising T1 | -0.007 | 0.006 | 0.264 | 0.968 | -0.005 | 0.007 | 0.434 | -0.024 | 0.020 | 0.236 |
| Forceps Minor FA T2 ~ Internalising T1 | -0.001 | 0.007 | 0.848 | 0.995 | -0.004 | 0.007 | 0.621 | 0.018 | 0.021 | 0.391 |
| Cingulate Gyrus FA T2 ~ Externalising T1 | 0.003 | 0.007 | 0.695 | 0.995 | 0.002 | 0.007 | 0.797 | 0.009 | 0.020 | 0.652 |
| Parahippocampal Cingulum FA T2 ~ Externalising T1 | -0.010 | 0.006 | 0.096 | 0.968 | -0.007 | 0.006 | 0.229 | -0.036 | 0.020 | 0.076 |
| Corticospinal Tract FA T2 ~ Externalising T1 | -0.013 | 0.006 | 0.031 | 0.968 | -0.014 | 0.006 | 0.027 | -0.004 | 0.019 | 0.841 |
| Anterior Thalamic Radiation FA T2 ~ Externalising T1 | -0.002 | 0.006 | 0.742 | 0.995 | 0.001 | 0.007 | 0.887 | -0.029 | 0.020 | 0.148 |
| Uncinate FA T2 ~ Externalising T1 | 0.001 | 0.006 | 0.919 | 0.995 | 0.002 | 0.007 | 0.724 | -0.011 | 0.018 | 0.527 |
| Inferior Longitudinal Fasciculus FA T2 ~ Externalising T1 | -0.006 | 0.006 | 0.318 | 0.968 | -0.005 | 0.007 | 0.501 | -0.017 | 0.017 | 0.310 |
| Inferior-fronto-occipital Fasciculus FA T2 ~ Externalising T1 | -0.009 | 0.006 | 0.148 | 0.968 | -0.009 | 0.006 | 0.174 | -0.008 | 0.016 | 0.620 |
| Superior Longitudinal Fasciculus FA T2 ~ Externalising T1 | -0.004 | 0.006 | 0.567 | 0.995 | -0.005 | 0.007 | 0.444 | 0.005 | 0.015 | 0.759 |

| | Meta-analysis | | | | ABCD | | | GenR | | |
|---|---|---|---|---|---|---|---|---|---|---|
| Forceps Major FA T2 ~ Externalising T1 | -0.005 | 0.006 | 0.402 | 0.995 | -0.002 | 0.007 | 0.710 | -0.031 | 0.020 | 0.126 |
| Forceps Minor FA T2 ~ Externalising T1 | 0.000 | 0.007 | 0.970 | 0.995 | 0.000 | 0.007 | 0.961 | -0.001 | 0.021 | 0.978 |
| ***Tract-based MD T1 → Psychiatric symptoms T2*** | | | | | | | | | | |
| Internalising T2 ~ Cingulate Gyrus MD T1 | 0.007 | 0.010 | 0.486 | 0.995 | 0.014 | 0.011 | 0.231 | -0.023 | 0.025 | 0.353 |
| Internalising T2 ~ Parahippocampal Cingulum MD T1 | 0.017 | 0.010 | 0.104 | 0.968 | 0.017 | 0.011 | 0.144 | 0.017 | 0.025 | 0.477 |
| Internalising T2 ~ Corticospinal Tract MD T1 | 0.014 | 0.010 | 0.173 | 0.968 | 0.019 | 0.011 | 0.100 | -0.007 | 0.024 | 0.767 |
| Internalising T2 ~ Anterior Thalamic Radiation MD T1 | 0.000 | 0.010 | 0.988 | 0.995 | 0.008 | 0.011 | 0.495 | -0.036 | 0.024 | 0.136 |
| Internalising T2 ~ Uncinate MD T1 | 0.014 | 0.010 | 0.169 | 0.968 | 0.019 | 0.011 | 0.088 | -0.010 | 0.024 | 0.684 |
| Internalising T2 ~ Inferior Longitudinal Fasciculus MD T1 | 0.016 | 0.010 | 0.118 | 0.968 | 0.013 | 0.011 | 0.272 | 0.035 | 0.025 | 0.172 |
| Internalising T2 ~ Inferior-fronto-occipital Fasciculus MD T1 | 0.015 | 0.010 | 0.156 | 0.968 | 0.017 | 0.011 | 0.133 | 0.003 | 0.025 | 0.900 |
| Internalising T2 ~ Superior Longitudinal Fasciculus MD T1 | 0.010 | 0.010 | 0.335 | 0.968 | 0.005 | 0.011 | 0.678 | 0.035 | 0.025 | 0.159 |
| Internalising T2 ~ Forceps Major MD T1 | 0.008 | 0.010 | 0.450 | 0.995 | 0.012 | 0.011 | 0.286 | -0.013 | 0.025 | 0.611 |
| Internalising T2 ~ Forceps Minor MD T1 | 0.012 | 0.010 | 0.249 | 0.968 | 0.015 | 0.011 | 0.199 | -0.001 | 0.024 | 0.976 |
| Externalising T2 ~ Cingulate Gyrus MD T1 | 0.005 | 0.010 | 0.634 | 0.995 | 0.009 | 0.011 | 0.392 | -0.019 | 0.025 | 0.437 |
| Externalising T2 ~ Parahippocampal Cingulum MD T1 | 0.018 | 0.010 | 0.068 | 0.968 | 0.017 | 0.011 | 0.124 | 0.026 | 0.025 | 0.299 |
| Externalising T2 ~ Corticospinal Tract MD T1 | 0.013 | 0.010 | 0.180 | 0.968 | 0.016 | 0.011 | 0.146 | 0.001 | 0.025 | 0.978 |
| Externalising T2 ~ Anterior Thalamic Radiation MD T1 | -0.002 | 0.010 | 0.841 | 0.995 | 0.005 | 0.011 | 0.648 | -0.038 | 0.024 | 0.123 |
| Externalising T2 ~ Uncinate MD T1 | 0.006 | 0.010 | 0.573 | 0.995 | 0.008 | 0.011 | 0.484 | -0.005 | 0.025 | 0.846 |
| Externalising T2 ~ Inferior Longitudinal Fasciculus MD T1 | -0.001 | 0.010 | 0.924 | 0.995 | -0.004 | 0.011 | 0.729 | 0.015 | 0.026 | 0.564 |
| Externalising T2 ~ Inferior-fronto-occipital Fasciculus MD T1 | 0.008 | 0.010 | 0.427 | 0.995 | 0.010 | 0.011 | 0.341 | -0.005 | 0.025 | 0.838 |
| Externalising T2 ~ Superior Longitudinal Fasciculus MD T1 | 0.000 | 0.010 | 0.995 | 0.995 | -0.004 | 0.011 | 0.739 | 0.019 | 0.025 | 0.444 |

| | Meta-analysis | | | | ABCD | | | GenR | | |
|---|---|---|---|---|---|---|---|---|---|---|
| Externalising T2 ~ Forceps Major MD T1 | 0.006 | 0.010 | 0.519 | 0.995 | 0.007 | 0.011 | 0.509 | 0.002 | 0.025 | 0.923 |
| Externalising T2 ~ Forceps Minor MD T1 | 0.005 | 0.010 | 0.645 | 0.995 | 0.006 | 0.011 | 0.584 | -0.003 | 0.025 | 0.918 |
| ***Psychiatric symptoms T1 → Tract-based MD T2*** | | | | | | | | | | |
| Cingulate Gyrus MD T2 ~ Internalising T1 | -0.008 | 0.007 | 0.258 | 0.968 | -0.003 | 0.008 | 0.717 | -0.049 | 0.021 | 0.021 |
| Parahippocampal Cingulum MD T2 ~ Internalising T1 | -0.005 | 0.008 | 0.485 | 0.995 | -0.003 | 0.008 | 0.715 | -0.022 | 0.022 | 0.309 |
| Corticospinal Tract MD T2 ~ Internalising T1 | -0.001 | 0.007 | 0.909 | 0.995 | 0.002 | 0.008 | 0.845 | -0.025 | 0.025 | 0.318 |
| Anterior Thalamic Radiation MD T2 ~ Internalising T1 | -0.004 | 0.007 | 0.565 | 0.995 | -0.005 | 0.008 | 0.491 | 0.004 | 0.021 | 0.833 |
| Uncinate MD T2 ~ Internalising T1 | 0.000 | 0.008 | 0.960 | 0.995 | -0.001 | 0.009 | 0.901 | 0.008 | 0.020 | 0.676 |
| Inferior Longitudinal Fasciculus MD T2 ~ Internalising T1 | 0.004 | 0.006 | 0.532 | 0.995 | 0.002 | 0.007 | 0.728 | 0.015 | 0.018 | 0.392 |
| Inferior-fronto-occipital Fasciculus MD T2 ~ Internalising T1 | 0.004 | 0.007 | 0.505 | 0.995 | 0.003 | 0.007 | 0.713 | 0.015 | 0.018 | 0.387 |
| Superior Longitudinal Fasciculus MD T2 ~ Internalising T1 | -0.001 | 0.007 | 0.893 | 0.995 | 0.003 | 0.008 | 0.668 | -0.018 | 0.016 | 0.252 |
| Forceps Major MD T2 ~ Internalising T1 | 0.007 | 0.007 | 0.329 | 0.968 | 0.002 | 0.007 | 0.771 | 0.039 | 0.020 | 0.048 |
| Forceps Minor MD T2 ~ Internalising T1 | -0.004 | 0.009 | 0.660 | 0.995 | -0.006 | 0.009 | 0.485 | 0.014 | 0.024 | 0.550 |
| Cingulate Gyrus MD T2 ~ Externalising T1 | -0.006 | 0.007 | 0.384 | 0.995 | -0.007 | 0.008 | 0.346 | 0.001 | 0.021 | 0.965 |
| Parahippocampal Cingulum MD T2 ~ Externalising T1 | -0.007 | 0.008 | 0.351 | 0.968 | -0.006 | 0.008 | 0.467 | -0.016 | 0.022 | 0.471 |
| Corticospinal Tract MD T2 ~ Externalising T1 | -0.002 | 0.007 | 0.799 | 0.995 | -0.001 | 0.008 | 0.862 | -0.007 | 0.025 | 0.768 |
| Anterior Thalamic Radiation MD T2 ~ Externalising T1 | -0.003 | 0.007 | 0.704 | 0.995 | -0.008 | 0.008 | 0.293 | 0.037 | 0.021 | 0.076 |
| Uncinate MD T2 ~ Externalising T1 | -0.009 | 0.008 | 0.254 | 0.968 | -0.016 | 0.009 | 0.058 | 0.030 | 0.020 | 0.128 |
| Inferior Longitudinal Fasciculus MD T2 ~ Externalising T1 | -0.003 | 0.006 | 0.676 | 0.995 | -0.006 | 0.007 | 0.329 | 0.025 | 0.018 | 0.154 |
| Inferior-fronto-occipital Fasciculus MD T2 ~ Externalising T1 | 0.001 | 0.007 | 0.871 | 0.995 | -0.006 | 0.007 | 0.417 | 0.042 | 0.018 | 0.016 |
| Superior Longitudinal Fasciculus MD T2 ~ Externalising T1 | -0.002 | 0.007 | 0.758 | 0.995 | 0.000 | 0.008 | 0.957 | -0.009 | 0.016 | 0.563 |

| | Meta-analysis | | | | ABCD | | | GenR | | |
|---|---|---|---|---|---|---|---|---|---|---|
| Forceps Major MD T2 ~ Externalising T1 | -0.003 | 0.007 | 0.635 | 0.995 | -0.008 | 0.007 | 0.308 | 0.027 | 0.020 | 0.175 |
| Forceps Minor MD T2 ~ Externalising T1 | -0.008 | 0.009 | 0.341 | 0.968 | -0.016 | 0.009 | 0.087 | 0.044 | 0.024 | 0.069 |

*Note*. ~ = regressed on; FA = fractional anisotropy; MD = mean diffusivity; *p* = *p*-value; SE = standard error; T = time-point.

**Supplementary Table 10.** Model fit comparison for models where sexes were restricted vs. allowed to vary in regressions

| Model | Df | AIC | BIC | Chisq | Chisq diff | Df diff | *p* |
|---|---|---|---|---|---|---|---|
| **The ABCD Study** | | | | | | | |
| *Internalizing problems* | | | | | | | |
| Internalizing FA (base) | 128 | 40466.56 | 46849.98 | 323.11 | NA | NA | NA |
| Internalizing FA (equal reg.) | 180 | 40402.40 | 46451.20 | 362.95 | 39.84 | 52 | 0.891 |
| Internalizing MD (base) | 128 | 34538.66 | 40922.08 | 298.19 | NA | NA | NA |
| Internalizing MD (equal reg.) | 180 | 34475.55 | 40524.36 | 339.08 | 40.89 | 52 | 0.867 |
| *Externalizing problems* | | | | | | | |
| Externalizing FA (base) | 128 | 40868.51 | 47251.93 | 342.77 | NA | NA | NA |
| Externalizing FA (equal reg.) | 180 | 40810.34 | 46859.14 | 388.60 | 45.83 | 52 | 0.714 |
| Externalizing MD (base) | 128 | 34943.33 | 41326.75 | 324.85 | NA | NA | NA |
| Externalizing MD (equal reg.) | 180 | 34884.27 | 40933.07 | 369.78 | 44.93 | 52 | 0.746 |

| Model | Df | AIC | BIC | Chisq | Chisq diff | Df diff | *p* |
|---|---|---|---|---|---|---|---|
| **The Generation R Study** | | | | | | | |
| *Internalizing problems* | | | | | | | |
| Internalizing FA (base) | 60 | 20552.83 | 21442.56 | 105.47 | NA | NA | NA |
| Internalizing FA (equal reg.) | 74 | 20554.54 | 21374.30 | 135.19 | 29.71 | 14 | 0.008 |
| Internalizing MD (base) | 60 | 21217.48 | 22107.22 | 133.48 | NA | NA | NA |
| Internalizing MD (equal reg.) | 74 | 21199.12 | 22018.88 | 143.12 | 9.64 | 14 | 0.788 |
| *Externalizing problems* | | | | | | | |
| Externalizing FA (base) | 60 | 20613.45 | 21503.18 | 116.49 | NA | NA | NA |
| Externalizing FA (equal reg.) | 74 | 20614.44 | 21434.19 | 145.48 | 28.99 | 14 | 0.010 |
| Externalizing MD (base) | 60 | 21275.23 | 22164.96 | 146.50 | NA | NA | NA |
| Externalizing MD (equal reg.) | 74 | 21255.21 | 22074.97 | 154.48 | 7.98 | 14 | 0.890 |

*Note.* AIC = Akaike's Information Criterion; BIC = Bayesian Information Criterion; Chisq = chi-square; Diff = difference; Df = degrees of freedom; NA = not applicable; reg = regression coefficients
